# Continuous indices to assess the phenotypic spectrum of kidney transplant rejection

**DOI:** 10.1101/2024.12.10.24318058

**Authors:** Thibaut Vaulet, Priyanka Koshy, Karolien Wellekens, Olivier Aubert, Charlotte Bottomley, Jasper Callemeyn, Evert Cleenders, Maarten Coemans, Lynn Cornell, Aiko P.J. de Vries, Gillian Divard, Marie-Paule Emonds, Sandrine Florquin, Mark Haas, Philip F. Halloran, Jesper Kers, Dirk Kuypers, Thangamani Muthukumar, Angelica Pagliazzi, Steven Salvatore, Olivier Thaunat, Surya V. Seshan, Elisabet Van Loon, Thomas Vanhoutte, Georg A Böhmig, Friedrich A. von Samson-Himmelstjerna, Michelle Willicombe, Aravind Cherukuri, Alexandre Loupy, Candice Roufosse, Maarten Naesens

## Abstract

The international Banff classification for kidney transplant pathology discretizes the rejection continuum into distinct diagnostic categories, introducing artificial dichotomization and threshold effects. To better reflect the underlying disease spectrum, we developed, in this cohort study, two novel indices for quantifying antibody-mediated (AMR) and T-cell mediated rejection (TCMR) from histological lesion scores, and calculated indices for overall activity and chronicity. These indices were evaluated in one derivation cohort and two independent validation cohorts, totaling 19,500 biopsies from 8,873 kidney transplant patients across 10 centers worldwide. The AMR, TCMR, and activity indices demonstrated hierarchical ordering between No rejection, intermediate and complete rejection histology. The chronicity index showed limited association with the major diagnostic categories. In the derivation cohort, the AMR and TCMR indices discriminated AMR from absence of AMR, and TCMR from absence of TCMR, with an AUC of 0.98 (95% confidence interval 0.97 to 0.98) and 0.99 (0.99 to 1.00) respectively). This excellent discrimination was confirmed in the validation cohorts. Those indices strictly confined intermediate phenotypes to a range of low index values and related to graft failure rates even within the diagnostic categories, thus reflecting the underlying rejection continuum. The four continuous indices offer an implementable and interpretable global evaluation of kidney transplant biopsy histology while eliminating the need for intermediate diagnostic categories and enable more probabilistic reasoning in the diagnostic approach to the spectrum of kidney transplant rejection.

## Introduction

Like many pathologic processes, kidney transplant rejection is a gradual and continuous process, more or less active depending on donor-recipient genetic disparity, highly heterogeneous immunologic responses to this genetic mismatch, and the efficacy of the immunosuppression prescribed to the recipients.^1,2^ The diagnosis of kidney transplant rejection currently relies on the Banff classification for kidney transplant pathology. This is an expert-based consensus framework for the diagnosis of allograft rejection and its subtypes, primarily antibody-mediated rejection (AMR) and T-cell mediated rejection (TCMR), characterized histologically by microvascular and tubulointerstitial inflammation, respectively.^3^ As such, this classification is essentially a discretization of the continuous rejection process into distinct diagnostic categories, which can be translated into complex decision trees like the Banff Automation System.^4^

Such classification introduces artificial cut-points that do not adequately reflect disease biology and disease spectrum. Categorization of diseases into non-overlapping classes aggregates highly diverse cases within singular diagnostic entities. Conversely, cases with very similar features but situated on opposite sides of a (consensus-based) diagnostic threshold are classified differently, while they are biologically equivalent.^5^ To partly mitigate this issue, many classifications and clinical guidelines adopt intermediate categories to bring more nuance.

Specific for kidney transplantation, the Banff classification introduced intermediate diagnostic categories such as “probable AMR” and “borderline for TCMR”, as well as acute/active, chronic-active, and chronic subcategories of AMR and TCMR, intended to reflect the temporal stage of disease. The clinical significance of such intermediate and temporal subcategories remains ambiguous and leads to recurring discussions on the choice of the boundaries between presence/absence of disease and different disease stages.^3,6^

More continuous quantification of the underlying disease process, closer to the actual pathological observation and to biological reality, could overcome the problems created by the artificial boundaries in typical disease classification models.^7–9^ In that regard, we recently developed activity^10^ and chronicity indices^11^ from Banff lesion scores of kidney transplant biopsies, representing the overall amount of inflammation and of chronicity observed in a kidney transplant biopsy, respectively. Others have developed mathematical models for quantifying and distinguishing AMR and TCMR.^12^ As acknowledged in the recent Banff consensus meetings, these scores hold promise for stratifying biopsies by disease severity, independently of the histological diagnosis.^3,6^ However, experts in the field highlighted that to date, no study has demonstrated added value of continuous indices for evaluation of kidney transplant pathology in comparison to the current rule-based classification.^6^

Therefore, we hypothesized that AMR- and TCMR-specific continuous indices derived from the routinely assessed histological lesion scores enable quantification of the global spectrum of kidney transplant rejection. Secondly, we hypothesized that the Banff diagnostic subcategories of rejection, as active/acute, chronic active, and chronic could be replaced by more continuous activity and chronicity indices calculated from the histological lesion scores, adding more detailed evaluation of disease stage and severity.

To test these hypotheses, we developed two novel, easily calculable continuous indices representing of the AMR and TCMR spectra, using a large derivation cohort. We validated these indices with two additional large multicentric validation cohorts by analyzing (1) their relationship with the Banff diagnostic categories and (2) their ability to discriminate different clinical outcome *within* these categories. Finally, we assessed and validated (3) the association of continuous activity and chronicity indices with graft failure rates, independent of the Banff (sub)categories. These four continuous indices, all derived from standard lesion scores, elegantly capture the full histological spectrum and severity of kidney transplant rejection, more comprehensively than the Banff (sub)categories.

## Results

### Patient and biopsy characteristics

The details of the population characteristics are reported in Table 1. After excluding patients without post-transplantation biopsy (N=77) and biopsies of inadequate quality (N=119), a total of 6,272 post-transplant biopsies from 1,814 patients were included in the derivation cohort. Validation was performed on two external cohorts: the EU validation cohort consisted of 11,043 biopsies from 5,898 transplants and the US validation cohort of 2,185 biopsies from 1,161 transplants.

**TABLE 1.** Demographic, clinical, and histological characteristics of the patients and biopsies included in the derivation and validation cohorts. BMI: body-mass index, eGFR: estimated glomerular filtration rate. SD: standard deviation. Percentages and statistical tests are derived from complete values.

| Cohort characteristics |  |  |  |  |  |
| --- | --- | --- | --- | --- | --- |
|  | Derivation cohort<br>N= 1814 | European validation Cohort<br>N= 5898 | P value* | US validation cohort<br>N=1161 | P value* |
| **Donor demographics** |  |  |  |  |  |
| Donor type |  |  | <0.001 |  | <0.001 |
| Donation after brain death, N (%) | 1315 (72.5) | 2900 (50.0) |  | 526 (45.6) |  |
| Donation after cardiac death, N (%) | 355 (19.6) | 1045 (18.0) |  | 169 (14.6) |  |
| Living donation, N (%) | 144 (7.9) | 1862 (32.0) |  | 460 (39.8) |  |
| Missing | 0 | 91 |  | 6 |  |
| Age (years), mean ± SD | 49.0 ± 14.4 | 52.2 ± 15.3 | <0.001 | 42.0 ± 13.9 | <0.001 |
| *missing* | 0 (0) | 61 (1.0) |  | 16 (1.1) |  |
| Male, N (%) | 985 (54.3) | 2931 (50.5) | 0.008 | 599 (51.6) | 0.19 |
| *missing* | 0 (0) | 97 (1.6) |  |  |  |
| **Recipient demographics** |  |  |  |  |  |
| Age (years), mean ± SD | 54.5 ± 12.8 | 50.7 ± 14.3 | <0.001 | 51.2 ± 14.7 | <0.001 |
| *Missing* | 0 (0) | 0 |  | 1 (0.1) |  |
| Male, N (%) | 1152 (63.5) | 3759 (63.7) | 0.88 | 691 (59.5) | 0.03 |
| Ethnicity<sup>#</sup> |  |  | <0.001 |  | <0.001 |
| Asian, N (%) | 14 (0.8) | 618 (31.0) |  | 40 (3.4) |  |
| Black, N (%) | 54 (3.0) | 283 (14.2) |  | 268 (23.1) |  |
| Others, N (%) | 1 (0.1) | 180 (9.0) |  | 49 (4.3) |  |
| White, N (%) | 1745 (96.2) | 912 (45.8) |  | 804 (69.3) |  |
| *missing* | 0 (0) | 3905 (66.2) |  | 0 (0) |  |
| BMI (kg/m<sup>2</sup>), mean ± SD<sup>‡</sup> | 25.6 ± 4.5 | 25.5 ± 4.8 | 0.68 | 27.4 ± 6.1 | <0.001 |
| *missing* |  | 892 (15.1) |  | 951 (81.9) |  |
| Pre-transplant donor-specific HLA antibodies, N (%) | 151 (8.3) | NA |  | NA |  |
| Repeat transplantation, N (%)<sup>¶</sup> | 273 (15.0) | 916 (18.8) |  | 277 (23.9) |  |
| *missing* | 0 (0) | 1028 (17.4) |  | 0 (0) |  |
| Cold ischemia time (hours), mean ± SD | 12.7 ± 6.0 | 13.0 ± 8.1 | 0.19 | 8.7 ± 7.5 | <0.001 |
| *missing* | 1 (0.1) | 929 (15.8) |  | 61 (5.3) |  |
| Total number of HLA A/B/DR mismatches, mean ± SD | 2.7 ± 1.3 | 3.3 ± 1.5 | <0.001 | 3.8 ± 1.6 | <0.001 |
| *missing* |  | 230 (3.9) |  | 9 (0.8) |  |
| N Graft failure | 255 (14.1) | 1009 (17.1) | 0.002 | 171 (14.7) | 0.578 |
| Biopsy characteristics | N= 6272 | N=11043 |  | N=2185 |  |
| Days since transplantation, median (interquartile range) | 362.0<br>(89.0-732.0) | 222.0<br>(83.0-630.0) | <0.001 | 168.0<br>(95.0-380.0) | <0.001 |
| Indication biopsies, N (%) | 1666 (26.6) | 6143 (55.8) | <0.001 | 854 (39.1) | <0.001 |
| *missing* |  | 37 (0.3%) |  |  |  |
| Banff 22 Categories |  |  |  |  |  |
| No rejection | 4327 (75.2) | 6738 (61.0) |  | 879 (40.2) |  |
| TCMR | 576 (9.2) | 1138 (10.3) |  | 629 (28.8) |  |
| Borderline TCMR | 537 (8.6) | 1083 (9.8) |  | 515 (23.6) |  |
| AMR | 343 (5.5) | 1175 (10.6) |  | 197 (9.0) |  |
| MV<sup>I</sup><sub>DSA-/C4d-</sub> | 278 (4.4) | 575 (5.2) |  | 101 (4.6) |  |
| PVAN | 161 (2.8) | 262 (2.4) |  | 76 (3.5) |  |
| Mixed rejection | 110 (1.8) | 287 (2.6) |  | 124 (5.7) |  |
| Probable AMR | 100 (1.6) | 210 (1.9) |  | 17 (0.8) |  |
| Unclassifiable<sup>†</sup> | 0 (0.0) | 705 (6.4) |  | 17 (0.8) |  |

\*In comparison with the derivation cohort, Student t-test and chi-square test, where appropriate.
<sup>#</sup>Ethnicity not available for centers Amsterdam, Leiden, Lyon, Paris and Vienna
<sup>‡</sup>BMI not available for University Hospital Schleswig-Holstein and University of Pittsburgh cohorts
<sup>¶</sup>Repeated transplantation information not included in University Hospital Schleswig-Holstein cohort
<sup>†</sup>Biopsies unclassifiable due to missing key Banff lesion score for definitive diagnosis.

All biopsies were reclassified according to the Banff 2022 classification^3^ using the individual Banff lesion scores and HLA-DSA status, into the following, potentially co-existing, diagnostic categories: AMR (containing active, chronic active, and chronic AMR); probable AMR (further referred to as “Probable AMR”); DSA-negative, C4d-negative microvascular inflammation (further referred to as “MVI_DSA-/C4d-_”); borderline/suspicious for TCMR (further referred to as “Borderline TCMR”); acute TCMR; and polyomavirus-associated nephropathy (further referred to as PVAN); normal or other changes as defined by the Banff category 6 (further referred to as “No rejection”). Cases with isolated v1 without tubulo-interstitial lesions (i0t0) were considered as No rejection instead of TCMR.^13^ In this study, “mixed rejection” strictly refers to concomitant presence of AMR and TCMR; “(Borderline) TCMR” refers to Borderline TCMR or TCMR/

The majority of transplants in the derivation cohort were from deceased donors (92.1%). In total, 273 (15.0%) transplants were repeat transplantations, and 151 (8.3%) had pretransplant HLA-DSA. Of all biopsies, 1,711 (27.3%) were performed for clinical indication, while the other 4,561 (72.7%) biopsies were performed per protocol. TCMR (9.2%) and Borderline TCMR (8.6%) were the most common rejection phenotypes. AMR occurred in 5.5%, MVI_DSA-/C4d-_ in 4.4%, Probable AMR in 1.6%, and mixed rejection in 1.8% of cases. A total of 4,327 (75.2%) biopsies did not meet the criteria for the diagnostic categories and were classified as No rejection biopsies. The details of the diagnostic categories in the validation cohorts are reported in Table 1.

During the period of observation, 248 (13.9%) kidney allografts failed in the derivation cohort. In the EU and US validation cohorts, 1,009 (17.1%) and 171 (14.7%) kidney allografts failed, respectively.

### Distribution of the continuous indices in relation to the Banff diagnostic (sub)categories

Based on the premise that AMR and TCMR constitute continuous disease spectra, AMR and TCMR can be represented by latent continuous variables instead of being regarded as strictly binary phenomena (present vs. absent). As such, we developed an AMR index and a TCMR index based on a latent variable approach, where the unobserved latent component is inferred from a set of relevant, observed Banff lesions, the binary diagnostic categories (AMR vs. no AMR, and TCMR vs. no TCMR) serving as dependent variable to guide the inference of the latent variable (Online Methods). The activity and chronicity indices were adapted from previously published models.^10,11^ The continuous indices included in the analyses, shown in Table 2, were calculated for all biopsies. Their distribution in the derivation cohort is visualized in Fig. 1.

**TABLE 2.**
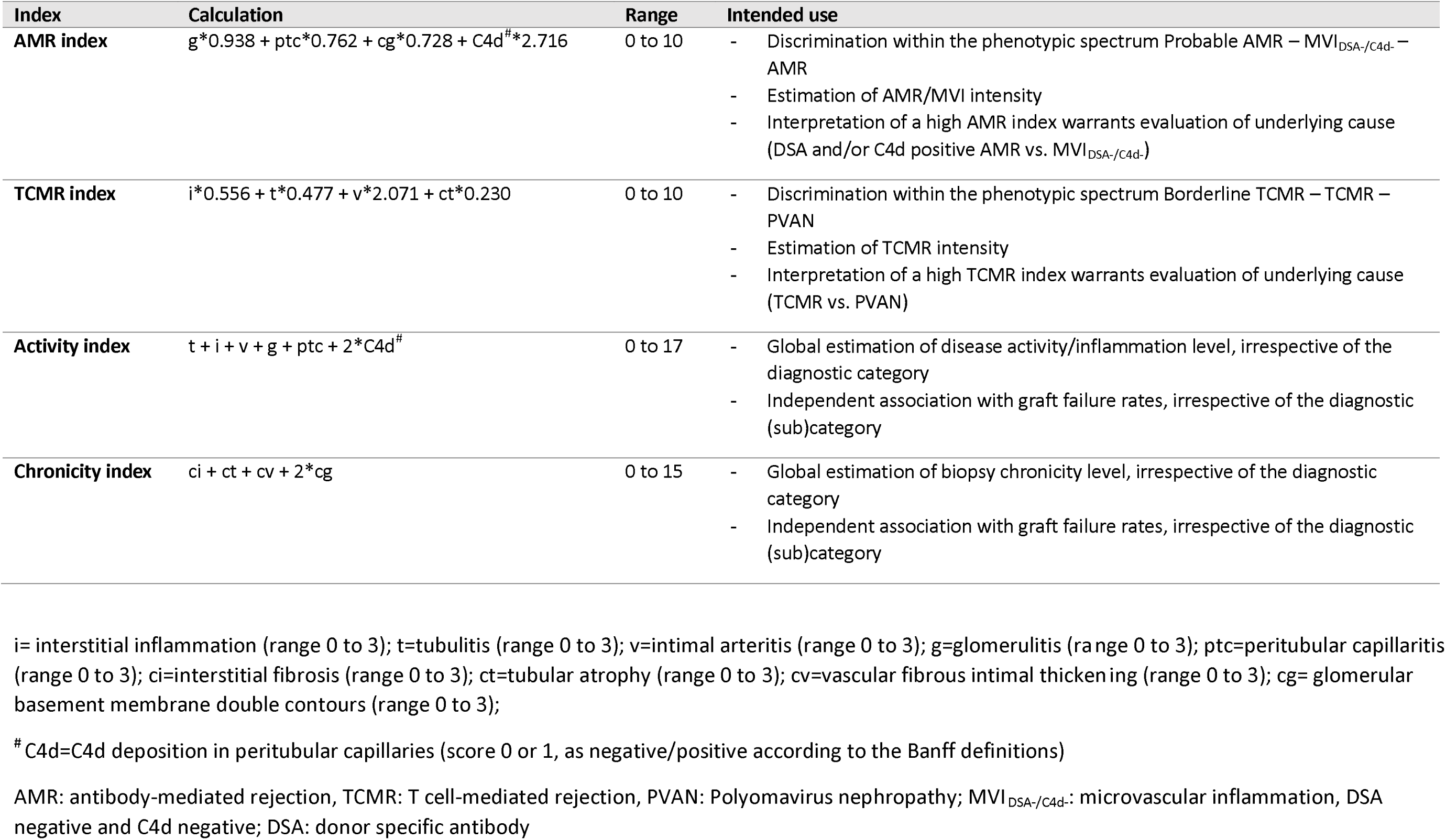
Overview of the continuous indices and their intended use.

**Figure 1.**
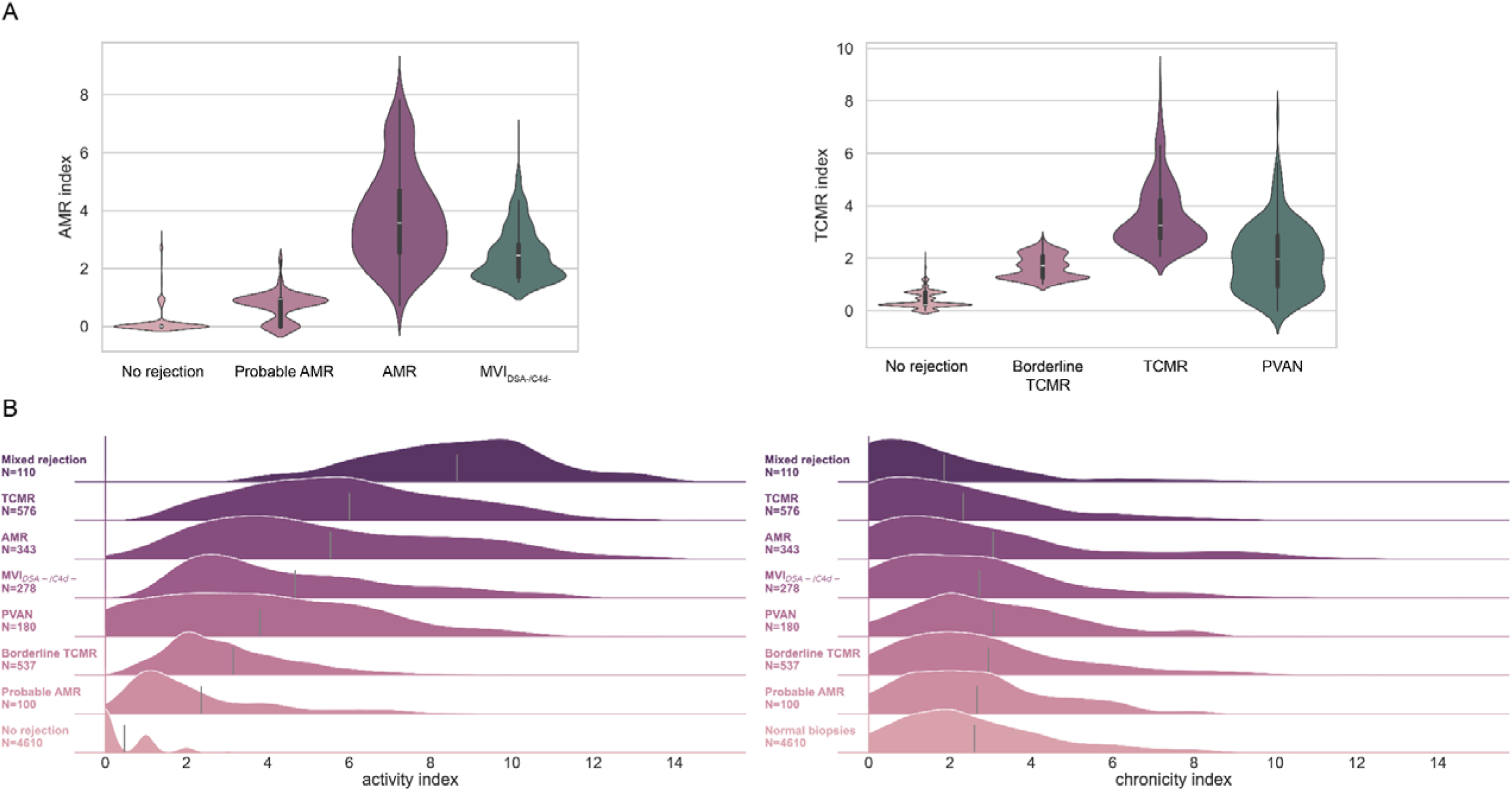
Distribution of the indices among the main Banff diagnoses in derivation cohort (N=6272 biopsies) **A.** Both AMR and TCMR indices demonstrated logical and hierarchical ordering between No rejection, intermediate (Probable AMR and Borderline TCMR) and complete rejection histology with, in some cases, significant overlap. **B.** Activity and chronicity indices, ordered by mean activity index per category (grey vertical line). The activity index demonstrated similar gradual and hierarchical ordering between No rejection biopsies and fully inflamed categories, with intermediate categories (Borderline TCMR, Probable AMR) in between. The chronicity index demonstrated low variability amongst the main diagnostic categories.

The AMR index gradually increased from No rejection to Probable AMR to AMR. MVI_DSA-/C4d-_ cases also exhibited high AMR indices. The TCMR index also demonstrated a gradual increase from No rejection to Borderline TCMR to TCMR. The TCMR index for PVAN cases was more broadly distributed and was significantly higher compared to No rejection (Student t-test: p<0.001), due to similar histological lesions as in TCMR in a subset of PVAN biopsies.

The Banff diagnostic categories were logically distributed in a two-dimensional space based on a combination of the AMR and the TCMR index (Fig. 2). AMR and MVI_DSA-/C4d-_ cases demonstrated significant overlap of in the AMR index. The highest AMR indices were observed to AMR cases, but this is inherent to the definition of MVI_DSA-/C4d-_ which requires C4d negativity. TCMR and Borderline TCMR cases fell within well-defined boundaries set by the TCMR index. Overall, the four indices were systematically higher in indication biopsies compared to protocol biopsies (Supplementary Fig. 1), albeit with significant overlap across the range, demonstrating that severe inflammation and chronic injury can occur subclinically, with stable graft function.

**Figure 2.**
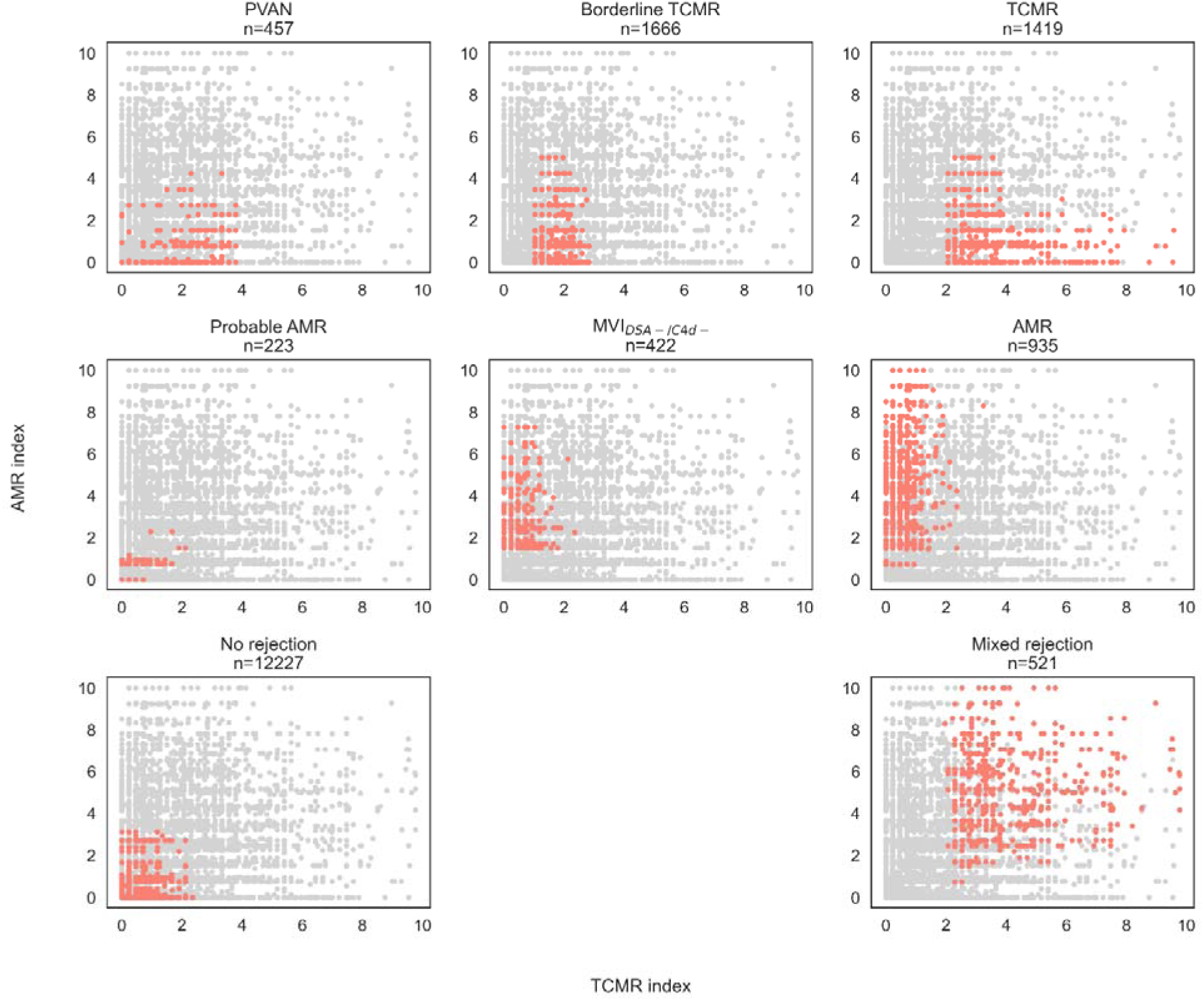
Distribution of the individual biopsies per diagnostic category (in red) based on the AMR (y axis) and TCMR indices (x axis) in all classifiable biopsies, N=18778. For visualization purposes, only the ‘pure’ phenotypes with no other concomitant diagnosis are displayed. No rejection biopsies are strictly characterized by both low AMR and low TCMR indices, whereas Mixed rejection cases have both high AMR and TCMR indices. High heterogeneity in Mixed rejection cases is however observed, resulting from the combination of various levels of intensity of the AMR and TCMR phenotypes within this diagnostic category. TCMR and Borderline TCMR cases are contained within clear boundaries of the TCMR index. Probable AMR cases are restricted to low AMR index. We note a significant overlap of the MVI_DSA-/C4d-_ cases with the AMR cases in terms of AMR index, although high values of AMR index are only affected to biopsies with AMR.

The activity index exhibited a gradual and hierarchical increase from No rejection to fully inflamed categories (AMR, MVI_DSA-/C4d-_, TCMR and Mixed rejection), with intermediate categories (Borderline TCMR, Probable AMR) in between (Fig. 1B). The chronicity index demonstrated low variability amongst the main diagnostic categories (Fig. 1B). Despite statistically different mean values of activity and chronicity indices between the acute/active, chronic-active, and chronic subcategories of AMR and TCMR, substantial overlap was observed in these indices across the subcategories (Supplementary Fig. 2). The latter analysis on the subcategories of TCMR was performed on a subset of the derivation cohort (N=1635) for which scoring of critical lesions of chronic active TCMR, namely ti and i-IFTA, were available. On another subset of 1254 biopsies with available lesions scores, the “i-IFTA” and “t-IFTA” scores correlated more strongly with the chronicity index (Spearman correlation: 0.40, p<0.001 and 0.43, p<0.001, respectively), than with the activity index (Spearman correlation: 0.10, p<0.001 and 0.26, p<0.001) and were therefore were not considered for inclusion in the activity index.

The distribution of the four continuous indices across the main diagnostic categories were similar in the validation cohorts compared with the derivation cohort (Supplementary Figs. 3 and 4), although the chronicity index was noticeably higher in both validation cohorts, with a larger spread across all main diagnostic categories. The correlation between the activity and the chronicity indices was also more pronounced in both the European and the US validation cohort (correlation coefficient [r] =0.18, P<0.001 and r=0.42, P<0.001, respectively), compared to a quasi-independence of the activity index and chronicity index (r=-0.08, P<0.001) in the derivation cohort.

### Discriminative performance of the AMR and TCMR indices for Banff diagnostic categories

To demonstrate that the novel AMR and TCMR indices maintain congruence with the clinically used Banff classification system of AMR and TCMR, we next evaluated the discriminative performance of the AMR and TCMR indices for the Banff diagnostic categories, analyzing the derivation and validation cohorts separately.

In the derivation cohort (Table 3), the AMR index discriminated presence of AMR from absence of AMR with an AUC of 0.98 (95% confidence interval [CI], 0.97 to 0.98). When MVI_DSA-/C4d-_ cases were combined with AMR, the AUC was 0.98 (95% CI, 0.98 to 0.99). The TCMR index demonstrated an AUC of 0.99 (95% CI, 0.99 to 1.00) for discriminating between the presence and absence of TCMR. This variable performed equally good if Borderline TCMR cases were considered as TCMR (AUC 0.99; 95% CI, 0.98 to 0.99). Compared to the other diagnostic categories, the TCMR index underperformed for the discrimination of PVAN, with an AUC of 0.81 (95% CI, 0.79-0.84), which can be explained by the absence of specific polyomavirus markers (e.g. SV40 staining results) in the TCMR index formulation. The activity index discriminated No rejection from all other categories with an AUC of 0.96 (95% CI, 0.95-0.96) and Mixed rejection from No Mixed rejection with an AUC of 0.98 (95% CI, 0.98-0.99), clearly outperforming both AMR and TCMR indices on those two tasks. Overall, the AUPRC, which is less impacted by significant class imbalance, demonstrated similar ordering of the variables performances among the different Banff categories as the AUC results (Supplementary Tables 1, 2 and 3).

**TABLE 3.** Discrimination performance as AUC with 95% CI of the acute indices (activity, AMR and TCMR indices) for relevant binary outcomes in all three cohorts. Note: biopsies unclassifiable (N=722) due to key lesion missing were excluded.

| Index | Discrimination | Derivation cohort<br>N=6272 | Validation EU<br>N=10338 | Validation US<br>N=2162 |
| --- | --- | --- | --- | --- |
| **AMR Index** | AMR vs No AMR | 0.98 (0.97-0.98) | 0.97 (0.96-0.97) | 0.96 (0.95-0.97) |
|  | MVI/AMR vs no MVI/no AMR | 0.98 (0.98-0.99) | 0.98 (0.98-0.98) | 0.97 (0.96-0.98) |
|  | Probable AMR/MVI/AMR vs no Probable AMR/MVI/AMR | 0.96 (0.95-0.96) | 0.96 (0.96-0.97) | 0.96 (0.95-0.97) |
| **TCMR Index** | TCMR vs No TCMR | 0.99 (0.99-1.00) | 0.99 (0.99-0.99) | 0.98 (0.97-0.98) |
|  | (Borderline) TCMR vs no (Borderline) TCMR | 0.99 (0.98-0.99) | 0.98 (0.98-0.98) | 0.95 (0.94-0.96) |
|  | PVAN vs No PVAN | 0.81 (0.79-0.84) | 0.79 (0.76-0.82) | 0.83 (0.80-0.86) |
| **Activity index** | Mixed rejection vs No Mixed rejection | 0.98 (0.98-0.99) | 0.98 (0.98-0.99) | 0.96 (0.95-0.97) |
|  | No rejection vs Any other category | 0.96 (0.95-0.96) | 0.97 (0.97-0.97) | 0.99 (0.99-0.99) |
|  | Any Rejection vs No rejection | 0.96 (0.95-0.96) | 0.95 (0.95-0.96) | 0.96 (0.95-0.97) |

As outlined in Table 3 and Supplementary Tables 1, 2 and 3, the performances of all indices were similar in both validation cohorts for all discrimination tasks.

Additionally, the discrimination performances of the three acute indices (activity, AMR and TCMR indices) were similar between indication and protocol biopsies (Supplementary Table 4). Net benefit of the AMR and TCMR indices was demonstrated for the discrimination of AMR and TCMR (Supplementary Results and Supplementary Fig. 5). The classification performances of the AMR and TCMR indices across their range are detailed in Supplementary Results: AMR and TCMR indices ≥3 had 99.2% and 99.3% specificity for respectively AMR/MVI_DSA-/C4d-_ and TCMR (Supplementary Tables 5, 6 and 7). All Probable AMR (N=327) and Borderline TCMR (N=2135) cases had respectively AMR and TCMR indices strictly inferior to 3, and strictly superior to 0 for Borderline TCMR cases (Supplementary Tables 8 and 9).

In the overall cohort, the AMR and TCMR indices yielded strata of increasing severity, indicated by the significant association of the AMR and TCMR indices with graft failure (Supplementary Fig. 6). Additionally, the indices effectively discriminated significantly different survival trends *within* the main Banff diagnostic categories. For instance, AMR cases with a higher AMR index exhibited a greater risk of graft failure than those with a lower AMR index. The same was true for MVI_DSA-/C4d-_ cases stratified by the AMR index, as well as for (Borderline) TCMR cases stratified by the TCMR index (Supplementary Fig. 7).

### Association of activity and chronicity indices with graft failure

To assess the relation between different disease stages and outcomes, we associated the activity and chronicity indices with long-term graft failure (Fig. 3; Supplementary Table 10).

**Figure 3.**
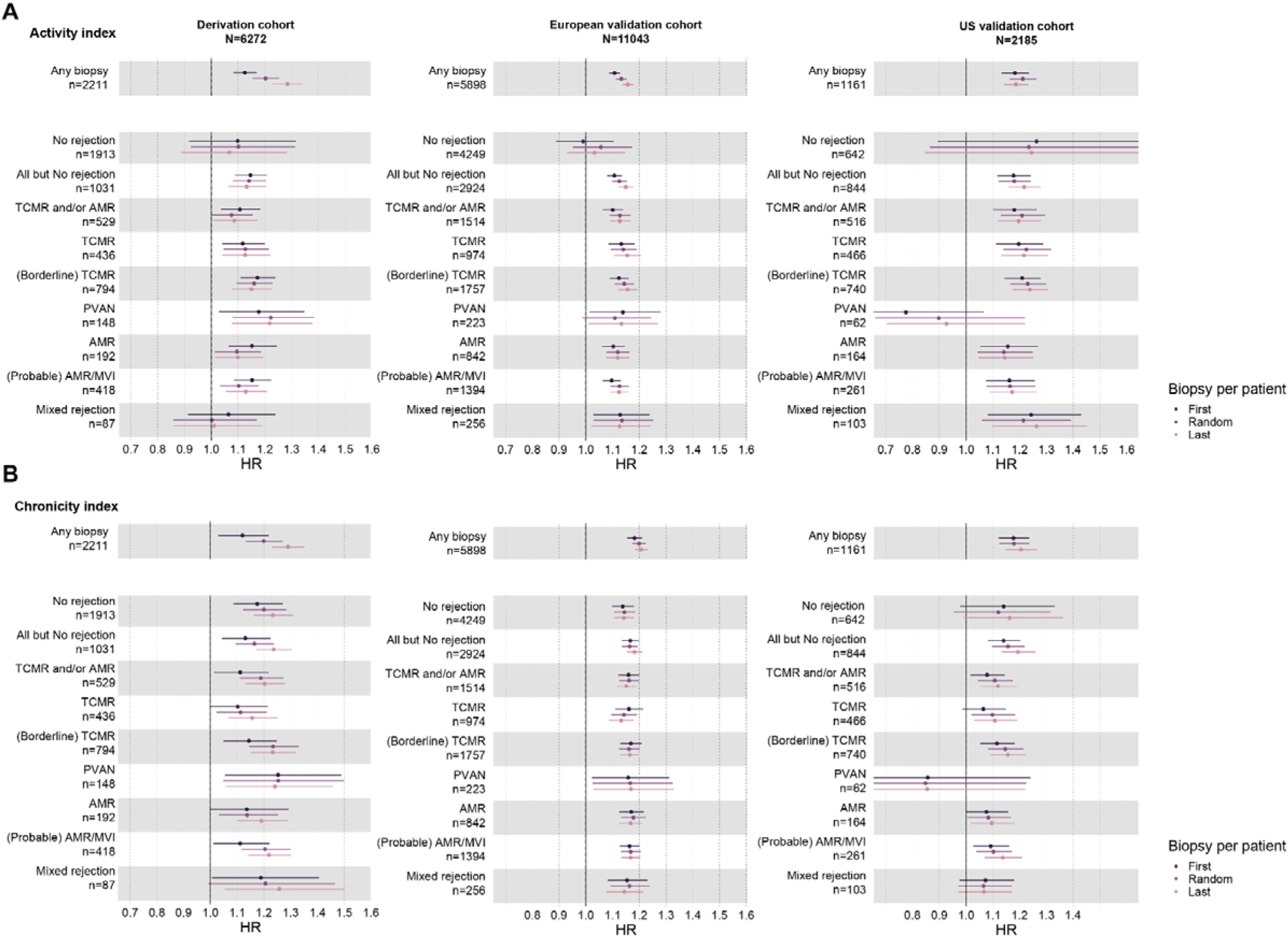
Association of the activity (panel A) and chronicity (panel B) indices with graft failure in the main Banff categories as hazard ratios [HR] (with 95% CI) adjusted for time post-transplantation in the three cohorts. HR refer to one-unit increase of the indices. The small n refers to number of unique patients (one biopsy per patient) per analysis.

In the derivation cohort, the continuous activity and chronicity indices associated with graft failure in Cox models adjusted for time post-transplantation, with an adjusted Hazard Ratio [HR] of 1.29 (95% CI, 1.23-1.34) and 1.29 (95% CI, 1.23-1.35), respectively (based on last biopsy per patient). Similar associations were observed in the two validation cohorts: the adjusted HR per unit increase of the activity index was 1.16 (95% CI 1.14-1.18) and of the chronicity index 1.21 (95% CI 1.18-1.23) in the European cohort; in the US validation cohort, these adjusted HRs were 1.19 (95% CI 1.14-1.23) and 1.21 (95% CI 1.15-1.26), respectively. In all three cohorts, the associations of activity and chronicity indices with graft failure were independent of the biopsy selected per patient. Of note, in a subset of the derivation cohort with available data (N=3724), replacing the “i” score in the activity with “ti” in the activity index did not alter the results (Supplementary Fig. 8).

The activity and chronicity indices demonstrated linear relations with graft failure rates (i.e. risk increasing at a constant rate with increased index, Supplementary Fig. 9). For each unit increase in the activity and the chronicity index, the relative hazard for graft failure increased by 15.0% (95% CI, 13.6 to 17.0%) and 20.6% (95% CI, 18.0 to 22.9%), respectively (computed on the whole cohort based on a random biopsy per patient).

These indices also maintained association with graft outcome *within* most of the main Banff-defined diagnostic categories. Only in No rejection and in Mixed rejection cases, which are less heterogeneous in terms of histological lesion scores, did the activity index not further stratify outcomes. Overall, similar associations of the indices with graft outcome were observed in both validation cohorts, except for PVAN in the US cohort. In contrast to the derivation cohort, the activity index even stratified outcomes within Mixed rejection cases in both validation cohorts.

The associations of the activity and chronicity indices with graft failure were observed both in indication and in protocol biopsies, except for the chronicity index which did not associate with graft failure in the subset of protocol biopsies classified as TCMR (Supplementary Fig. 10).

Both activity and chronicity indices remained associated with graft failure within the AMR active, chronic-active, and chronic subcategories (Supplementary Fig. 11, Supplementary Table 11). Similar analysis on the TCMR index could not be performed due to low prevalence of chronic active TCMR in the derivation subset.

### Discussion

This study shows that only four dimensions – four continuous measures of kidney transplant rejection derived from standard histological lesion scores – capture the full histological spectrum and severity of kidney transplant rejection, while closely aligning with the current Banff classification. The AMR and TCMR indices directly reflect the two distinct yet potentially concurrent inflammation patterns of kidney allografts: microvascular versus tubulo-interstitial respectively. The activity and chronicity indices provide a global evaluation of the level of inflammation and level of chronic damage, respectively. The AMR and TCMR indices effectively discriminate between their corresponding Banff-defined rejection categories and constitute simple yet meaningful proxies for the continuous rejection processes, also when TCMR and AMR processes are present simultaneously in mixed rejection phenotypes. The AMR and TCMR indices correlate with graft outcomes even within the Banff diagnostic categories. The activity index helps distinguish between rejection and no rejection and assesses inflammation severity, the latter being associated with graft failure independent of the underlying disease, and even within the Banff AMR acute/active, chronic-active, and chronic subcategories. The chronicity index offers prognostic information independent of the diagnostic category and of disease activity. For every one-unit increase in the activity and the chronicity indices, we observed a linear increase of respectively 15.0% and 20.6% in the risk of graft failure.

Not only are the AMR and TCMR indices discriminating full rejection phenotypes providing a detailed view on the disease spectrum, but these indices are also increased in cases borderline/suspicious for TCMR and with diagnosis of probable AMR, albeit below the index values of fully developed phenotypes. This supports their clinical interpretation of such cases as intermediate phenotypes rather than distinct diagnostic categories.^14,15^ Moreover, the relationship between AMR and TCMR indices and graft outcomes, even within these intermediate phenotypes, highlights the additional information regarding disease severity that the continuous scores provide compared to the Banff categories.

At the other side of the spectrum, the large histological heterogeneity of the mixed rejection cases is particularly apparent when studying the combination of AMR and TCMR indices: next to high AMR/high TCMR indices, combinations such as low AMR/high TCMR indices or high AMR/low TCMR indices are all labelled similarly as Mixed rejection. The AMR and TCMR indices provide a more detailed evaluation of the relative contributions of the AMR versus TCMR components in such mixed cases.

While there is generally strong concordance between the continuous AMR and TCMR indices and the Banff classification, their relationship with other phenotypes warrants some further discussion. The histology of MVI_DSA-/C4d-_ resembles that of AMR, with glomerulitis and/or peritubular capillaritis, resulting in high AMR scores for this phenotype as well. The only difference in the Banff classification of these cases and Banff-defined AMR is the underlying disease cause is not directly detected in routine histology. Similarly, the histology of PVAN often resembles TCMR, exhibiting tubulo-interstitial inflammation.^16^ It is therefore no surprise that the TCMR index is often high in such PVAN cases, as would be the cases for other diseases with a tubulointerstitial nephritis phenotype from other causes (e.g. drug toxicity). The underperformance of the TCMR index to discriminate PVAN cases is explained by the lack of polyomavirus-specific markers in the index formulation (e.g. SV40 positive staining) and the considerable heterogeneity in PVAN presentations which can range from non-inflamed to heavily inflamed. Overall, the associations of the AMR (microvascular) and TCMR (tubulo-interstitial) indices with MVI_DSA-/C4d-_ and PVAN, respectively, underscore the necessity of interpreting the histological picture in conjunction with additional testing for underlying causes. Specifically, HLA-DSA testing is crucial for distinguishing AMR from MVI_DSA-/C4d-_ and SV40 staining (and peripheral blood PCR) is essential for differentiating between TCMR and PVAN diagnosis.

The AMR and TCMR indices offer a detailed view on the disease spectrum, avoiding the arbitrary thresholds between intermediate diagnostic categories like Borderline TCMR and Probable AMR. Yet, as we demonstrated that the relation between the diagnostic categories and the indices is very strong, reporting on the continuous variables in e.g. pathology reports can be easily done in parallel to the discretized diagnostic categories, without affecting their interpretation. In the vast majority of cases, our data confirm that the indices align well with the diagnostic categories. Discrepancies are mostly found in those expected cases where the Banff system potentially over-emphasizes the relevance of certain markers (e.g. HLA-DSA positive cases with minimal inflammation considered as AMR; underestimation of the relevance of t3+i1 or i3+t1 in cases classified as Borderline TCMR). From this perspective, it can even be expected that increasing experience with the continuous variables calculated from Banff lesion scores, and perhaps in the future with biopsy-based transcript analyses, will eventually also lead to refinement of the Banff definitions for the diagnostic categories per se.

In this study, the activity and chronicity indices were associated with graft failure risk and showed high heterogeneity and relationship to outcomes within the AMR subcategories (active, chronic-active, chronic). This indicates that these subcategories do not fully capture disease stage, as intended. Replacing subcategories with activity and chronicity indices in biopsy reports could improve clinical decision-making and offer more granularity for clinical trials. Highly active disease could benefit from treatment with anti-inflammatory or targeted therapies aimed at the immune activation and its underlying etiology. More chronic disease presentations with lesser activity could be less reversible by such etiological therapies. Although intuitive, the clinical value and benefit of grading activity and chronicity alongside diagnostic categories should be tested in prospective trials.

Our activity and chronicity indices were constructed based on previous mathematical modelling, with further refinement based on discussions within the Banff Working Group. Other algorithms could however be considered. For instance, Haas et al. have proposed an alternative activity index (g+ptc+v+C4d), specific for AMR cases.^17^ However, they could not demonstrate an association with graft failure in this specific setting. In addition, it could be considered to include more recently defined Banff lesion scores in the models, such as ti, i-IFTA and t-IFTA. We however demonstrated that replacing the Banff i score for ti does not affect the results, and that i-IFTA and t-IFTA are highly collinear with chronic lesions, potentially confounding activity, and chronicity if these parameters would be included in an activity index. Moreover, the lack of large-enough datasets in which these lesions have been evaluated systematically hinders robust statistical analysis of the added value of including these lesions in the models. Further improvements to the activity and chronicity score definitions proposed herein should be tested in future studies.

This study has some limitations. The continuous indices require histological Banff lesion scores which remain inherently pathologist dependent. Emerging techniques like whole slide image analysis could improve reproducibility of lesion scoring and derived indices. Similarly, the Banff diagnostic categories, used as ground truth here, have evolved during the course of the study and the Banff 2022 classification has been attributed retrospectively. Also in the future, the Banff classification may evolve, though the distinction between essentially tubulo-interstitial disease and microvascular disease has been stable over two decades, reflecting distinct biological mechanisms. The latter supports long-term validity of the TCMR (tubulo-interstitial) and AMR (microvascular) indices, also with future updates of the Banff classification. Notwithstanding, future refinements of the indices should be explored. Therapeutic approaches differ between centers and treatment options and protocols have evolved over the course of the study, potentially affecting the relationship between histology and outcomes. In this study, stratifications were for illustration purpose only, and continuous variables should ideally not be discretized.^18^ Also grouping biopsies into broader rejection categories may mask heterogeneity. Finally, although our study is not a true multicenter study by design, the robustness of the indices was confirmed through validation in diverse and heterogeneous datasets, reflecting real-world clinical data and ensuring the generalizability of our findings beyond the controlled conditions of a traditional multicenter study.

In conclusion, we showed that continuous indices calculated from Banff lesion scores offer a straightforward and interpretable evaluation of kidney transplant biopsy histology, generalizable across centers in Europe and the US. The AMR and TCMR indices differentiate histological phenotypes and quantify the rejection spectrum, potentially removing the need for intermediate categories like Borderline TCMR or Probable AMR. Additionally, the activity and chronicity indices hold significant clinical relevance, as each one-unit increase is associated with an overall 15-20% higher risk of graft failure. Overall, these indices offer a more nuanced understanding of the continuous rejection process and support probabilistic reasoning in the diagnosis of kidney transplant rejection.

## Online Methods

### Study population and clinical data

This multicenter observational cohort study included 8,977 adult kidney transplant recipients from 10 European and United States transplant centers.

The derivation cohort consisted of all adult kidney transplantations performed at the University Hospitals Leuven, Belgium, between March 2004 and May 2021 (N=1,891; ClinicalTrials.gov ID NCT06505200). The transplants were performed with negative complement-dependent cytotoxicity crossmatches. Combined transplants and kidney transplants following a different solid organ transplant were excluded. The standard immunosuppressive maintenance protocol consisted of tacrolimus, mycophenolate, and corticosteroids^19^. Post-transplant biopsies were conducted based upon medical indication (indication biopsies at time of graft dysfunction) or as part of an established follow-up protocol (protocol biopsies), primarily at 3-, 12-, and 24-months post-transplantation. All clinical data for the derivation cohort were prospectively gathered in electronic health records. Data inclusion concluded on June 11^th^, 2022. Clinical data included donor demographics (type of donor, age, sex, diabetes), recipient demographics (age, sex, ethnicity, body mass index), and transplant characteristics (donor-specific HLA antibodies, HLA antigen mismatches, cold ischemia time, graft failure, mortality). Ethnic background was gathered to facilitate discussions on the generalizability of the findings. Graft failure was defined as return to dialysis or repeat transplantation. In case of death with a functioning graft, we censored graft failure at the time of death. Patients were censored at the time of their last known follow-up if no event occurred or if they were lost to follow-up.

We performed validation in two external cohorts: a European validation cohort consisting of 5,898 transplants and a US validation cohort of 1,161 transplants. In these validation cohorts, we obtained the same clinical and biological data as those collected for the derivation cohort according to routine clinical care, in accordance with local and national regulations and submitted to the central research database in Leuven (Supplementary Methods).

The study was approved by the Ethics Committee Research of the University Hospitals Leuven (S64006), with data transfer agreements with the contributing centers. No written informed consent was required for inclusion in this observational cohort study. The research was performed in accordance with the Declaration of Helsinki and the Guidelines for Good Clinical Practice.

### Kidney transplant biopsy evaluation

All allograft biopsies in the derivation cohort were prospectively catalogued in the electronic health records of the patients. Pre- and posttransplant human leukocyte antigens donor-specific antibodies (HLA-DSA) status were monitored in one histocompatibility laboratory (HILA – Red Cross Flanders) as reported previously for this cohort.^20^

In both the derivation and the validation cohorts, all transplant biopsies were evaluated by semiquantitative scoring of Banff lesion scores according Reference Guide to the Banff 2022 classification (https://banfffoundation.org/central-repository-for-banff-classification-resources-3/). Biopsies not reaching the threshold for minimal sample requirement were excluded^21^.

### Development of continuous histological indices

#### AMR and TCMR indices

The AMR and TCMR indices were based on a latent-variable approach, where the unobserved latent component is inferred from the observed Banff lesion scores, the binary diagnostic categories (AMR vs. no AMR, and TCMR vs. no TCMR) serving as dependent variable to guide the inference of the latent variable. More specifically, an ordinal regression model from the Python statsmodels package^1^ was trained to model the relationship between the observed Banff lesions and the binary diagnostic outcome. In these models, the binary outcome is treated as a threshold response to the continuous latent variable, rather than the probability of being present or absent, as in a logistic regression setting. Additionally, these models produced non-negative continuous latent variables, ideal to develop scoring systems. Based on the Banff classification, we established a list of potential variable candidates for the indices: g, ptc, v, C4d, v, i, t, ci, ct, cv and cg. As part of the modeling process, a feature selection step was performed to remove lesion scores that did not significantly contribute to the latent variable: backward selection was used to sequentially eliminate the Banff lesions that were not significant (at a p-value threshold of 0.05) in the latent variable. Finally, for ease of clinical use, we scaled both latent variables to the [0,10] interval (based on the combination of highest lesion scores) to produce the final AMR and TCMR indices. All biopsies were considered independent. Model fitting was conducted using maximum likelihood estimation, and performance was assessed with AUC for the relevant diagnostic categories, ensuring that the inferred indices reliably reflected the association between the lesions and the diagnostic outcome.

#### Activity and chronicity indices

Previously, we built activity and chronicity indices, as the sum of the active and chronic Banff lesion scores reweighted with the normalized coefficient’s *z* score of univariate Cox models for graft failure, and scaled to the unit interval (from 0 to 1; https://rejectionclass.eu.pythonanywhere.com)^10,11^:

⍰ Activity index= 0.049*t + 0.061*i + 0.066*v + 0.062*g + 0.062*ptc + 0.050*thrombi +0.052*C4d + 0.169*DSA
⍰ Chronicity index= 0.100*cg+ 0.074*ci+ 0.050*ct+ 0.038*cv+ 0.061*mm+ 0.052*ah+ 0.049*gs

Here, we simplified the activity index as t+i+v+g+ptc+2xC4d (score 0-17), where C4d is a binary indicator (0 or 1) of C4d positivity, which avoids the issue of heterogeneity in C4d staining methods between centers. In addition, HLA-DSA as a non-histological parameter was dropped out of the original formulation after discussion within the Banff Activity and Chronicity Indices Working Group (LC, MH, MN, SS). For the chronicity index, the simplification of the initial formula led to the same chronicity index as defined by Haas et al.^4^, i.e. 2xcg+ci+ct+cv (score 0-15), cg having a higher association with graft outcome, as reported in our original chronicity index formulation.^3^ To maximize clinical implementation across centers, we retained only the most clearly defined, reproducible and frequently reported Banff lesions for construction of the continuous indices. However, also more recently defined Banff lesions, representing both activity and chronicity (total inflammation score “ti”; interstitial inflammation in scarred area “i-IFTA”; tubulitis in scarred area “t-IFTA”) were considered in updated formulas.

### Missing data

Missing data were imputed with Multiple Imputation by Chained Equations (MICE)^22^, using predictive mean matching to generate 20 imputed datasets. Data visualizations, not directly aggregable according to Rubin’s rule, were produced based on the average linear predictors from those 20 sets. Validations datasets were imputed independently of the derivation cohort. Data were assumed to be missing at random. The diagnostic categories were used as additional proxy binary variables to guide the imputation. The proportion of missing histological lesion scores per cohort are reported in Supplementary Table 12.

### Outcome parameters

The main outcomes of interest were the discriminative performances of the four continuous indices (activity/chronicity index; AMR/TCMR index) for Banff-defined diagnostic categories and the association of these indices with graft outcome.

### Statistical analysis

We followed the STROBE guidelines for reporting on observational studies (Supplementary Appendix).

The discriminative performance of the indices was assessed based on the presence versus absence of Banff-defined phenotypes, with intermediate phenotypes first considered in the no disease category and subsequently taken together with complete phenotypes. Unclassifiable biopsies (due to key Banff lesion missing in validation cohorts) were excluded from discrimination analyses. The Area Under the ROC curves (AUC) and Precision-Recall curve (AUPRC) were used as discrimination metrics. We reported raw AUPRC and AURPC adjusted for the positive rate of each category in the cohort for comparison purposes. The biopsies were considered independent. To evaluate the clinical potential of an index beyond the rank-based metrics, Decision Curved Analysis (DCA) was applied.^23^ DCA evaluates the classification over a broad range of discretization thresholds and compare the benefit to two extremes decision policies^23^: classifying *none* of the biopsies as AMR (or TCMR) or *every* biopsy as AMR (or TCMR) classifying all biopsies as *positive* or all biopsies as *negative* for a given phenotype. The AMR and TCMR indices were compared to AMR and TCMR regression models (models 2 without HLA-DSA and Panel Reactive Antibody) as proposed earlier^12^ in terms of Net Benefit. The indices were transformed to the same numerical range by using individual logistic regression models trained on a given binary outcome (e.g. AMR versus No AMR). The correlation between a pair of indices was assessed with the Repeated Measures Correlation.^24^

Graft failure was defined as the return to dialysis or retransplantation. Patients were censored at the last follow-up date or at time of death. The association with graft failure rates was evaluated with Cox models for all indices. As a sensitivity analysis, either the first, last or a random biopsy per patient were selected to build the models in case of repeated biopsies. The overall association is reported with hazard ratios (HR), adjusted for time post-transplantation.

The main analyses were performed under R version 4.2.0 (R Foundation for Statistical Computing). The following packages were used: the MICE package^25^ for multiple imputation, the RMS package^26^ for discrimination performance, the survival package^27^ for survival analyses, the dcurves package^28^ for DCA and rmcorr package^24^ for the correlation adjusted for repeated measures. The figures were plotted with the ggplot2^29^, survminer^30^ and ggforestplot^31^ packages. The remaining figures were produced under Python 3.9.7. using matlplotlib^32^ and seaborn^33^. The latent variable models were produced using statsmodels 0.13.5.^34^

## Author contributions

TV and MN designed the study. AP, EC, EVL, JC, KW, MN, PK, TV, were involved in data collection and curation. AC, AdV, AL, CB, CR, FvSS, GAB, GD, JK, MC, OA, OT, SS, SVS, TM provided the validation data. TV performed all analysis with input from MN. TV and MN wrote the manuscript. All co-authors revised the manuscript.

## Data availability

The data from the derivation cohort is available upon motivated request to the corresponding author.

