## Supplementary Methods for "Continuous indices to assess the phenotypic spectrum of kidney transplant rejection"

**Description of the validation cohorts**

**Amsterdam University Medical Center (AMC)**

The Amsterdam University Medical Center validation cohort consisted of adult kidney transplant recipients followed at the location Academic Medical Center, Amsterdam, The Netherlands, who underwent an allograft biopsy procedure performed between 2000 and 2019, according to routine clinical practice. All kidney biopsies were retrospectively re-assessed according to the latest Banff iteration (Banff '22) by a single academic renal pathologist. The data collection was approved by the IRB of Amsterdam UMC (registration #19.260). All kidney transplant biopsies were performed either for indication or per protocol (in the context of randomized clinical trials at 6, 12 or 24 months post-transplantation).

**Leiden University Medical Center (LUMC)**

The Leiden University Medical Center validation cohort consisted of adult kidney transplant recipients followed at the LUMC, Leiden, The Netherlands, who underwent an allograft biopsy procedure performed between 2011 and 2024, according to routine clinical practice. All kidney biopsies were retrospectively re-assessed according to the latest Banff iteration (Banff '22) by a single academic renal pathologist. The data collection was approved by the IRB of Leiden-Den Haag-Delft (registration # W2020.031) All kidney transplant biopsies were performed either for indication or per protocol (in the context of randomized clinical trials at 6, or 12 months post-transplantation).

**Necker Hospital Paris**

The Paris validation cohort consisted of adult kidney transplant recipients followed at Necker Hospital, Paris, France who underwent an allograft biopsy procedure performed between March 2009 and October 2019, according to routine clinical practice. The data collection was approved by the IRB of Paris Transplant Institute (registration LMN2019X01). All kidney transplant biopsies were performed either for indication or per protocol, as part of the routine follow-up at 3 months and 12 months post transplantation.

**Lyon**

The Lyon validation cohort consisted of adult kidney transplant recipients followed at Lyon University Hospitals who underwent an allograft biopsy procedure performed between January 2007 and December 2015. The data collection was approved by the IRB Comité de protection des personnes Sud-Est I (registration #2020-A02918-31). All kidney transplant biopsies were performed according to routine clinical practice, either for indication or per protocol, as part of the routine follow-up at 3 months and 12 months post transplantation.

**Imperial College London**

The Imperial College London validation cohort consisted of adult kidney transplant recipients followed at Imperial College Healthcare NHS Trust who underwent an allograft biopsy procedure performed between 05.01.2007 and 26.03.2024, according to routine clinical practice. Human samples and data were obtained from the Imperial College Healthcare Tissue & Biobank (ICHTB), under project R14094. ICHTB is supported by the National Institute for Health Research (NIHR) Biomedical Research Centre based at Imperial College Healthcare NHS Trust and Imperial College London. ICHTB is approved by Wales REC3 to release human material for research (22/WA/2836). All kidney transplant biopsies were performed either for indication or per protocol, as part of the routine follow-up at 3 months and 12 months post transplantation.

**University Hospital Schleswig-Holstein (UKSH)**

The University Hospital Schleswig-Holstein (UKSH) validation cohort consisted of adult kidney transplant recipients followed at UKSH, Kiel Germany, who underwent an allograft biopsy procedure performed between 06 Apr 2018 and 07 Dec 2022, according to routine clinical practice. The data collection was approved by the IRB of the Medical Faculty of the Christian-Albrechts-University Kiel (registration #B278/16). All kidney transplant biopsies were performed on indication.

**Medical University Vienna**

The Medical University Vienna validation cohort consisted of adult kidney transplant recipients primarily followed at the Medical University of Vienna, Vienna, Austria, who underwent an allograft biopsy procedure performed between 1. Jan 2013 and 1. Sept 2023, according to routine clinical practice. The data collection was approved by the IRB of the Medical University of Vienna (EK-Nr. 267/2011). All kidney transplant biopsies were performed for indication or per protocol, as part of the routine follow-up at 3 months, 12 months and/or 3 years post transplantation.

**Weill Cornell Medicine**

The Weill Cornell Medicine (WCM) validation cohort consisted of adult kidney transplant recipients followed at the NewYork Presbyterian Hospital - Weill Cornell Medical Center, New York, who underwent an allograft biopsy procedure performed between January 2019 and April 2022, according to routine clinical practice. Biopsies were done for clinically indicated reasons. The data collection was approved by the WCM Institutional Review Board (#1404015008).

**University of Pittsburgh**

The University of Pittsburgh validation cohort consisted of adult kidney transplant recipients followed at the University of Pittsburgh Medical Center, Pittsburgh, Pennsylvania, who underwent an allograft biopsy procedure performed between the 1st of January 2013 and the 31st of December 2018, according to routine clinical practice. The data collection was approved by the IRB of the University of Pittsburgh School of Medicine (#19080302). All kidney transplant biopsies were performed either for indication or per protocol, as part of the routine follow-up at 3 months and 12 months post transplantation.

**SUPPLEMENTARY RESULTS**

**Net benefit for classification**

In all three cohorts, the AMR and TCMR indices demonstrated greater net benefit over most of the threshold range for the discrimination of AMR and TCMR respectively, compared to previously published[1] models and to the activity index (Supplementary Figure 5).

*Classification performance of the AMR and TCMR indices*

The classification performances of the AMR and TCMR indices for Banff-defined AMR and TCMR indicated that all Banff-defined TCMR cases (N=2343) had a TCMR index greater than 1.9 (sensitivity 100%) whereas biopsies with a TCMR index ≥3 (N=1293) were most often classified as TCMR according to the Banff classification (positive predictive value of 91.8% and specificity of 99.3%) (Supplementary Table 5). For AMR, biopsies with an AMR index ≥3 (N=1304) were classified as AMR with a specificity of 96.9% (Supplementary Table 6). Only 23 AMR cases (1.3% of AMR cases) had an AMR index less than 1; these cases were all chronic AMR with cg1 and HLA-DSA positive, but without any other AMR lesions (g0,ptc0,C4d0). This resulted in a sensitivity of 98.7% at an AMR threshold of 1. Additionally, all biopsies with AMR index ≥3 (N=1704) were almost all AMR or $MVI_{DSA\text{-}C4d\text{-}}$ (positive predicted value of 93.1% and specificity of 99.2%) (Supplementary Table 7). For intermediate phenotypes, all Probable AMR (N=327) had AMR indices strictly inferior to 3 and all Borderline TCMR (N=2135) had a TCMR index strictly superior to 0 and strictly inferior to 3 (Supplementary Tables 8 and 9).

**References**

1. Sikosana, M. L. N., Reeve, J., Madill-Thomsen, K. S., Halloran, P. F. & Investigators, the I. Using Regression Equations to Enhance Interpretation of Histology Lesions of Kidney Transplant Rejection. *Transplantation* 10.1097/TP.0000000000004783 doi:10.1097/TP.0000000000004783.
