## Supplementary for "Continuous indices to assess the phenotypic spectrum of kidney transplant rejection"

**TABLE S1. Discrimination performances of the AMR index and the activity index on AMR, $MVI_{DSA-/C4d}$ and Probable AMR outcomes, per cohort.**

pAMR: Probable AMR, MVI: $MVI_{DSA-/C4d}$

| | | Derivation cohort N=6272 | | | European validation cohort N=10339 | | | US validation cohort N=2162 | | |
|---|---|---|---|---|---|---|---|---|---|---|
| **Index** | **metric** | **AMR vs No AMR** | **MVI/AMR vs no MVI/no AMR** | **pAMR/MVI/AMR vs no pAMR/MVI/AMR** | **AMR vs No AMR** | **MVI/AMR vs no MVI/no AMR** | **pAMR/MVI/AMR vs no pAMR/MVI/AMR** | **AMR vs No AMR** | **MVI/AMR vs no MVI/no AMR** | **pAMR/MVI/AMR vs no pAMR/MVI/AMR** |
| **AMR index** | AUC | 0.98 (0.97-0.98) | 0.98 (0.98-0.99) | 0.96 (0.95-0.96) | 0.97 (0.96-0.97) | 0.98 (0.98-0.98) | 0.96 (0.96-0.97) | 0.96 (0.95-0.97) | 0.97 (0.96-0.98) | 0.96 (0.95-0.97) |
| **activity index** | AUC | 0.91 (0.90-0.92) | 0.92 (0.92-0.93) | 0.90 (0.89-0.91) | 0.90 (0.89-0.90) | 0.92 (0.91-0.92) | 0.90 (0.89-0.91) | 0.90 (0.88-0.92) | 0.91 (0.89-0.92) | 0.90 (0.88-0.92) |
| **AMR index** | Brier | 0.03 | 0.04 | 0.05 | 0.04 | 0.05 | 0.06 | 0.04 | 0.05 | 0.06 |
| **activity index** | Brier | 0.04 | 0.06 | 0.07 | 0.08 | 0.09 | 0.1 | 0.06 | 0.07 | 0.08 |
| **AMR index** | AUPRC | 0.74 | 0.86 | 0.8 | 0.8 | 0.91 | 0.87 | 0.7 | 0.84 | 0.83 |
| **activity index** | AUPRC | 0.4 | 0.57 | 0.56 | 0.51 | 0.7 | 0.69 | 0.54 | 0.67 | 0.68 |
| **AMR index** | AUPRC adj. | 13.58 | 8.64 | 6.99 | 7.06 | 5.35 | 4.61 | 7.73 | 6.14 | 5.7 |
| **activity index** | AUPRC adj. | 7.31 | 5.77 | 4.89 | 4.48 | 4.13 | 3.64 | 5.94 | 4.87 | 4.65 |

**TABLE S2. Discrimination performances of the TCMR index and the activity index, on TCMR, Borderline TCMR and PVAN outcomes, per cohort.**

(b)TCMR: Borderline TCMR or TCMR

| | | Derivation cohort N=6272 | | | European validation cohort N=10339 | | | US validation cohort N=2162 | | |
|---|---|---|---|---|---|---|---|---|---|---|
| **Index** | **metric** | **TCMR vs No TCMR** | **(b)TCMR Vs No (b)TCMR** | **PVAN vs No PVAN** | **TCMR vs No TCMR** | **(b)TCMR Vs No (b)TCMR** | **PVAN vs No PVAN** | **TCMR vs No TCMR** | **(b)TCMR Vs No (b)TCMR** | **PVAN vs No PVAN** |
| **TCMR index** | AUC | 0.99 (0.99-1.00) | 0.99 (0.98-0.99) | 0.81 (0.79-0.84) | 0.99 (0.99-0.99) | 0.98 (0.98-0.98) | 0.79 (0.76-0.82) | 0.98 (0.97-0.98) | 0.95 (0.94-0.96) | 0.83 (0.8-0.86) |
| **activity index** | AUC | 0.96 (0.96-0.97) | 0.94 (0.94-0.95) | 0.79 (0.76-0.82) | 0.96 (0.95-0.96) | 0.94 (0.94-0.95) | 0.72 (0.69-0.75) | 0.95 (0.94-0.96) | 0.91 (0.90-0.93) | 0.80 (0.77-0.83) |
| **TCMR index** | Brier | 0.02 | 0.04 | 0.03 | 0.02 | 0.05 | 0.02 | 0.05 | 0.07 | 0.03 |
| **activity index** | Brier | 0.04 | 0.08 | 0.03 | 0.05 | 0.09 | 0.02 | 0.09 | 0.12 | 0.03 |
| **TCMR index** | AUPRC | 0.94 | 0.92 | 0.09 | 0.93 | 0.91 | 0.07 | 0.91 | 0.92 | 0.1 |
| **activity index** | AUPRC | 0.72 | 0.77 | 0.09 | 0.72 | 0.8 | 0.05 | 0.85 | 0.89 | 0.08 |
| **TCMR index** | AUPRC adj. | 10.22 | 5.19 | 3.02 | 8.49 | 4.25 | 2.79 | 3.14 | 1.74 | 2.92 |
| **activity index** | AUPRC adj. | 7.86 | 4.33 | 3.04 | 6.57 | 3.7 | 1.91 | 2.92 | 1.68 | 2.32 |

**TABLE S3. Discrimination performances of the AMR, TCMR index and the activity index for Any abnormal biopsy, Mixed rejection and TCMR and/or AMR outcomes, per cohort.**

| | | Derivation cohort N=6272 | | | European validation cohort N=10339 | | | US validation cohort N=2162 | | |
|---|---|---|---|---|---|---|---|---|---|---|
| Index | | No rejection vs Any other category | Mixed rejection vs No mixed rejection | TCMR/AMR vs no Rejection | No rejection vs Any other category | Mixed rejection vs No mixed rejection | TCMR/AMR vs no Rejection | No rejection vs Any other category | Mixed rejection vs No mixed rejection | TCMR/AMR vs no Rejection |
| **TCMR index** | AUC | 0.88 (0.87-0.89) | 0.97 (0.96-0.97) | 0.87 (0.85-0.89) | 0.87 (0.86-0.88) | 0.97 (0.96-0.97) | 0.99 (0.98-0.99) | 0.82 (0.81-0.84) | 0.92 (0.91-0.94) | 0.94 (0.93-0.95) |
| **AMR index** | AUC | 0.76 (0.75-0.78) | 0.98 (0.97-0.98) | 0.81 (0.79-0.82) | 0.81 (0.8-0.82) | 0.95 (0.95-0.96) | 0.75 (0.74-0.77) | 0.86 (0.85-0.87) | 0.95 (0.94-0.96) | 0.81 (0.79-0.83) |
| **activity index** | AUC | 0.96 (0.95-0.96) | 0.98 (0.98-0.99) | 0.96 (0.95-0.96) | 0.97 (0.97-0.97) | 0.98 (0.98-0.99) | 0.99 (0.99-0.99) | 0.95 (0.95-0.96) | 0.96 (0.95-0.97) | 0.96 (0.95-0.97) |
| **TCMR index** | Brier | 0.08 | 0.02 | 0.05 | 0.11 | 0.02 | 0.03 | 0.1 | 0.04 | 0.07 |
| **AMR index** | Brier | 0.14 | 0.01 | 0.08 | 0.15 | 0.02 | 0.18 | 0.09 | 0.04 | 0.15 |
| **activity index** | Brier | 0.06 | 0.01 | 0.05 | 0.06 | 0.02 | 0.02 | 0.07 | 0.03 | 0.08 |
| **TCMR index** | AUPRC | 0.92 | 0.25 | 0.77 | 0.88 | 0.35 | 0.96 | 0.71 | 0.4 | 0.89 |
| **AMR index** | AUPRC | 0.86 | 0.39 | 0.59 | 0.84 | 0.3 | 0.59 | 0.71 | 0.46 | 0.71 |
| **activity index** | AUPRC | 0.98 | 0.53 | 0.79 | 0.98 | 0.62 | 0.99 | 0.85 | 0.63 | 0.9 |
| **TCMR index** | AUPRC adj | 1.25 | 13.97 | 5.98 | 1.36 | 12.61 | 2.38 | 3.63 | 6.98 | 2.74 |
| **AMR index** | AUPRC adj | 1.17 | 22.23 | 4.57 | 1.29 | 10.75 | 1.45 | 3.64 | 8.1 | 2.21 |
| **activity index** | AUPRC adj | 1.33 | 30.27 | 6.13 | 1.5 | 22.51 | 2.44 | 4.32 | 10.98 | 2.7 |

**TABLE S4 Sensitivity analyses- discrimination in AUC within Indication and Protocol biopsies.**

Biopsies without any known indication/protocol status were excluded. pAMR: Probable AMR, MVI: $MVI_{DSA-/C4d-}$ , (b)TCMR: Borderline TCMR or TCMR

| Index | Discrimination | Derivation cohort N=6272 | | Validation EU N=9107 | |
|---|---|---|---|---|---|
| | | Protocol N=4561 | Indication N=1711 | Protocol N=3390 | Indication N=5717 |
| AMR Index | AMR vs No AMR | 0.98 (0.97-0.98) | 0.97 (0.96-0.98) | 0.97 (0.96-0.98) | 0.96 (0.95-0.96) |
| | MVI/AMR vs no MVI/no AMR | 0.99 (0.98-0.99) | 0.97 (0.97-0.98) | 0.97 (0.97-0.98) | 0.98 (0.97-0.98) |
| | pAMR/MVI/AMR vs no pAMR/MVI/AMR | 0.96 (0.96-0.97) | 0.94 (0.92-0.95) | 0.95 (0.94-0.96) | 0.96 (0.95-0.96) |
| TCMR Index | TCMR vs No TCMR | 0.99 (0.99-1.00) | 0.99 (0.99-0.99) | 1.00 (0.99-1.00) | 0.99 (0.99-0.99) |
| | (b)TCMR vs no (b)TCMR | 0.99 (0.98-0.99) | 0.98 (0.97-0.99) | 0.99 (0.99-0.99) | 0.97 (0.97-0.98) |
| | PVAN vs No PVAN | 0.84 (0.81-0.88) | 0.74 (0.69-0.78) | 0.74 (0.61-0.88) | 0.75 (0.72-0.78) |
| Activity Index | Mixed rejection vs No Mixed rejection | 0.99 (0.99-1.00) | 0.96 (0.95-0.97) | 1.00 (0.99-1.00) | 0.97 (0.97-0.98) |
| | No rejection vs Any other category | 0.95 (0.94-0.96) | 0.97 (0.96-0.97) | 0.96 (0.96-0.97) | 0.97 (0.96-0.97) |
| | AMR/TCMR vs no rejection | 0.96 (0.95-0.96) | 0.94 (0.93-0.95) | 0.96 (0.95-0.97) | 0.94 (0.94-0.95) |

**TABLE S5 Classification metrics for arbitrary cut-off thresholds of the TCMR index for the classification of TCMR cases.** Based on all classifiable biopsies (N=18778 biopsies). The cut-off are interpreted as “index being greater or equal to the threshold”.

tp: true positive, tn: true negative, fp: false positive, fn: false negative, ppv: positive predictive value, npv: negative predictive value

| Threshold TCMR index | tp | tn | fp | fn | sensitivity | specificity | ppv | npv | accuracy |
|---|---|---|---|---|---|---|---|---|---|
| **0** | 2343 | 0 | 16435 | 0 | 1.000 | 0.000 | 0.125 | - | 0.125 |
| **1** | 2343 | 13228 | 3207 | 0 | 1.000 | 0.805 | 0.422 | 1.000 | 0.829 |
| **2** | 2342 | 15705 | 730 | 1 | 1.000 | 0.956 | 0.762 | 1.000 | 0.961 |
| **3** | 1293 | 16319 | 116 | 1050 | 0.552 | 0.993 | 0.918 | 0.940 | 0.938 |
| **4** | 573 | 16435 | 0 | 1770 | 0.245 | 1.000 | 1.000 | 0.903 | 0.906 |
| **5** | 342 | 16435 | 0 | 2001 | 0.146 | 1.000 | 1.000 | 0.891 | 0.893 |
| **6** | 180 | 16435 | 0 | 2163 | 0.077 | 1.000 | 1.000 | 0.884 | 0.885 |
| **7** | 94 | 16435 | 0 | 2249 | 0.040 | 1.000 | 1.000 | 0.880 | 0.880 |
| **8** | 32 | 16435 | 0 | 2311 | 0.014 | 1.000 | 1.000 | 0.877 | 0.877 |
| **9** | 19 | 16435 | 0 | 2324 | 0.008 | 1.000 | 1.000 | 0.876 | 0.876 |
| **10** | 0 | 16435 | 0 | 2343 | 0.000 | 1.000 | - | 0.875 | 0.875 |

**TABLE S6 Classification metrics for arbitrary cut-off thresholds of the AMR index for the classification of AMR, cases.** Based on all classifiable biopsies (N=18778 biopsies). The cut-off are interpreted as “index being greater or equal to the threshold”.

tp: true positive, tn: true negative, fp: false positive, fn: false negative, ppv: positive predictive value, npv: negative predictive value

| Threshold AMR index | tp | tn | fp | fn | sensitivity | specificity | ppv | npv | accuracy |
|---|---|---|---|---|---|---|---|---|---|
| **0** | 1715 | 0 | 17063 | 0 | 1.000 | 0.000 | 0.091 | - | 0.091 |
| **1** | 1692 | 14696 | 2367 | 23 | 0.987 | 0.861 | 0.417 | 0.998 | 0.873 |
| **2** | 1515 | 15472 | 1591 | 200 | 0.883 | 0.907 | 0.488 | 0.987 | 0.905 |
| **3** | 1304 | 16537 | 526 | 411 | 0.760 | 0.969 | 0.713 | 0.976 | 0.950 |
| **4** | 986 | 16763 | 300 | 729 | 0.575 | 0.982 | 0.767 | 0.958 | 0.945 |
| **5** | 714 | 16933 | 130 | 1001 | 0.416 | 0.992 | 0.846 | 0.944 | 0.940 |
| **6** | 413 | 17022 | 41 | 1302 | 0.241 | 0.998 | 0.910 | 0.929 | 0.928 |
| **7** | 249 | 17049 | 14 | 1466 | 0.145 | 0.999 | 0.947 | 0.921 | 0.921 |
| **8** | 120 | 17063 | 0 | 1595 | 0.070 | 1.000 | 1.000 | 0.915 | 0.915 |
| **9** | 70 | 17063 | 0 | 1645 | 0.041 | 1.000 | 1.000 | 0.912 | 0.912 |
| **10** | 28 | 17063 | 0 | 1687 | 0.016 | 1.000 | 1.000 | 0.910 | 0.910 |

**TABLE S7 Classification metrics for arbitrary cut-off thresholds of the AMR index for the classification of AMR/MVI$_{DSA-/C4d}$ cases.** Based on all classifiable biopsies (N=18778 biopsies). The cut-off are interpreted as "index being greater or equal to the threshold".

tp: true positive, tn: true negative, fp: false positive, fn: false negative, ppv: positive predictive value, npv: negative predictive value

| Threshold AMR index | tp | tn | fp | fn | sensitivity | specificity | ppv | npv | accuracy |
|---|---|---|---|---|---|---|---|---|---|
| **0** | 2669 | 0 | 16109 | 0 | 1.000 | 0.000 | 0.142 | - | 0.142 |
| **1** | 2646 | 14696 | 1413 | 23 | 0.991 | 0.912 | 0.652 | 0.998 | 0.924 |
| **2** | 2161 | 15164 | 945 | 508 | 0.810 | 0.941 | 0.696 | 0.968 | 0.923 |
| **3** | 1704 | 15983 | 126 | 965 | 0.638 | 0.992 | 0.931 | 0.943 | 0.942 |
| **4** | 1232 | 16055 | 54 | 1437 | 0.462 | 0.997 | 0.958 | 0.918 | 0.921 |
| **5** | 826 | 16091 | 18 | 1843 | 0.309 | 0.999 | 0.979 | 0.897 | 0.901 |
| **6** | 454 | 16109 | 0 | 2215 | 0.170 | 1.000 | 1.000 | 0.879 | 0.882 |
| **7** | 263 | 16109 | 0 | 2406 | 0.099 | 1.000 | 1.000 | 0.870 | 0.872 |
| **8** | 120 | 16109 | 0 | 2549 | 0.045 | 1.000 | 1.000 | 0.863 | 0.864 |
| **9** | 70 | 16109 | 0 | 2599 | 0.026 | 1.000 | 1.000 | 0.861 | 0.862 |
| **10** | 28 | 16109 | 0 | 2641 | 0.010 | 1.000 | 1.000 | 0.859 | 0.859 |

**TABLE S8 Classification metrics for arbitrary cut-off thresholds of the AMR index for the classification of Probable AMR cases.** Based on all classifiable biopsies (N=18778 biopsies). The cut-off are interpreted as “index being greater or equal to the threshold”.

tp: true positive, tn: true negative, fp: false positive, fn: false negative, ppv: positive predictive value, npv: negative predictive value

| Threshold AMR index | tp | tn | fp | fn | sensitivity | specificity | ppv | npv | accuracy |
|---|---|---|---|---|---|---|---|---|---|
| **0** | 327 | 0 | 18451 | 0 | 1.000 | 0.000 | 0.017 | - | 0.017 |
| **1** | 31 | 14423 | 4028 | 296 | 0.095 | 0.782 | 0.008 | 0.980 | 0.770 |
| **2** | 13 | 15358 | 3093 | 314 | 0.040 | 0.832 | 0.004 | 0.980 | 0.819 |
| **3** | 0 | 16621 | 1830 | 327 | 0.000 | 0.901 | 0.000 | 0.981 | 0.885 |
| **4** | 0 | 17165 | 1286 | 327 | 0.000 | 0.930 | 0.000 | 0.981 | 0.914 |
| **5** | 0 | 17607 | 844 | 327 | 0.000 | 0.954 | 0.000 | 0.982 | 0.938 |
| **6** | 0 | 17997 | 454 | 327 | 0.000 | 0.975 | 0.000 | 0.982 | 0.958 |
| **7** | 0 | 18188 | 263 | 327 | 0.000 | 0.986 | 0.000 | 0.982 | 0.969 |
| **8** | 0 | 18331 | 120 | 327 | 0.000 | 0.993 | 0.000 | 0.982 | 0.976 |
| **9** | 0 | 18381 | 70 | 327 | 0.000 | 0.996 | 0.000 | 0.983 | 0.979 |
| **10** | 0 | 18423 | 28 | 327 | 0.000 | 0.998 | 0.000 | 0.983 | 0.981 |

**TABLE S9 Classification metrics for arbitrary cut-off thresholds of the TCMR index for the classification of Borderline TCMR cases.** Based on all classifiable biopsies (N=18778 biopsies). The cut-off are interpreted as “index being greater or equal to the threshold”.

tp: True positive, tn: true negative, fp: false positive, fn: false negative, ppv: positive predictive value, npv: negative predictive value

| Threshold TCMR index | tp | tn | fp | fn | sensitivity | specificity | ppv | npv | accuracy |
|---|---|---|---|---|---|---|---|---|---|
| **0** | 2135 | 0 | 16643 | 0 | 1.000 | 0.000 | 0.114 | - | 0.114 |
| **1** | 2134 | 13228 | 3416 | 0 | 1.000 | 0.795 | 0.385 | 1.000 | 0.818 |
| **2** | 430 | 14001 | 2642 | 1705 | 0.201 | 0.841 | 0.140 | 0.891 | 0.769 |
| **3** | 0 | 15234 | 1409 | 2135 | 0.000 | 0.915 | 0.000 | 0.877 | 0.811 |
| **4** | 0 | 16070 | 573 | 2135 | 0.000 | 0.966 | 0.000 | 0.883 | 0.856 |
| **5** | 0 | 16301 | 342 | 2135 | 0.000 | 0.979 | 0.000 | 0.884 | 0.868 |
| **6** | 0 | 16463 | 180 | 2135 | 0.000 | 0.989 | 0.000 | 0.885 | 0.877 |
| **7** | 0 | 16549 | 94 | 2135 | 0.000 | 0.994 | 0.000 | 0.886 | 0.881 |
| **8** | 0 | 16611 | 32 | 2135 | 0.000 | 0.998 | 0.000 | 0.886 | 0.885 |
| **9** | 0 | 16624 | 19 | 2135 | 0.000 | 0.999 | 0.000 | 0.886 | 0.885 |
| **10** | 0 | 16643 | 0 | 2135 | 0.000 | 1.000 | - | 0.886 | 0.886 |

**Table S10 Hazard ratios (with 95% confidence interval) of the activity and chronicity indices in the three cohorts; adjusted for time post-transplantation.**

| ACTIVITY INDEX | Derivation cohort N=6272 | | | European validation cohort N=11043 | | | US validation cohort N=2185 | | |
|---|---|---|---|---|---|---|---|---|---|
| | First biopsy | Random biopsy | Last biopsy | First biopsy | Random biopsy | Last biopsy | First biopsy | Random biopsy | Last biopsy |
| All biopsies | 1.13 (1.08-1.17) | 1.20 (1.15-1.25) | 1.29 (1.23-1.34) | 1.11 (1.09-1.13) | 1.13 (1.11-1.15) | 1.16 (1.14-1.18) | 1.18 (1.13-1.23) | 1.21 (1.16-1.26) | 1.19 (1.14-1.23) |
| No rejection | 1.10 (0.92-1.32) | 1.08 (0.90-1.30) | 1.07 (0.89-1.28) | 0.99 (0.89-1.10) | 1.06 (0.95-1.17) | 1.03 (0.93-1.15) | 1.26 (0.89-1.78) | 1.23 (0.86-1.76) | 1.24 (0.85-1.83) |
| All but No rejection | 1.15 (1.09-1.21) | 1.14 (1.08-1.20) | 1.13 (1.07-1.20) | 1.11 (1.08-1.13) | 1.13 (1.10-1.15) | 1.15 (1.12-1.18) | 1.18 (1.12-1.24) | 1.18 (1.12-1.24) | 1.22 (1.16-1.28) |
| AMR and/or TCMR | 1.11 (1.04-1.18) | 1.08 (1.00-1.16) | 1.09 (1.01-1.17) | 1.10 (1.06-1.14) | 1.13 (1.09-1.17) | 1.13 (1.09-1.17) | 1.18 (1.10-1.26) | 1.21 (1.13-1.29) | 1.20 (1.12-1.28) |
| TCMR | 1.12 (1.04-1.20) | 1.13 (1.05-1.22) | 1.13 (1.04-1.22) | 1.13 (1.09-1.18) | 1.14 (1.09-1.19) | 1.16 (1.11-1.21) | 1.20 (1.11-1.29) | 1.23 (1.14-1.32) | 1.22 (1.13-1.31) |
| (Borderline) TCMR | 1.17 (1.11-1.24) | 1.16 (1.09-1.23) | 1.15 (1.08-1.23) | 1.12 (1.09-1.16) | 1.14 (1.11-1.18) | 1.16 (1.12-1.19) | 1.21 (1.14-1.28) | 1.23 (1.17-1.30) | 1.24 (1.17-1.31) |
| PVAN | 1.18 (1.03-1.35) | 1.22 (1.08-1.39) | 1.22 (1.08-1.38) | 1.14 (1.01-1.28) | 1.11 (0.99-1.24) | 1.13 (1.01-1.27) | 0.78 (0.56-1.07) | 0.90 (0.66-1.22) | 0.93 (0.70-1.22) |
| AMR | 1.15 (1.07-1.25) | 1.10 (1.01-1.19) | 1.10 (1.01-1.19) | 1.10 (1.06-1.15) | 1.12 (1.08-1.16) | 1.12 (1.08-1.16) | 1.16 (1.05-1.27) | 1.14 (1.04-1.25) | 1.14 (1.05-1.25) |
| Probable AMR /MVI/AMR | 1.15 (1.09-1.22) | 1.10 (1.03-1.18) | 1.13 (1.06-1.21) | 1.10 (1.06-1.13) | 1.13 (1.09-1.16) | 1.12 (1.09-1.16) | 1.16 (1.07-1.26) | 1.16 (1.08-1.26) | 1.17 (1.09-1.26) |
| Mixed rejection | 1.06 (0.91-1.24) | 1.00 (0.86-1.17) | 1.01 (0.86-1.19) | 1.13 (1.03-1.24) | 1.14 (1.03-1.25) | 1.13 (1.02-1.24) | 1.24 (1.08-1.43) | 1.21 (1.06-1.39) | 1.26 (1.10-1.45) |

| CHRONICITY INDEX | Derivation cohort N=6272 | | | European validation cohort N=11043 | | | US validation cohort N=2185 | | |
|---|---|---|---|---|---|---|---|---|---|
| | First biopsy | Random biopsy | Last biopsy | First biopsy | Random biopsy | Last biopsy | First biopsy | Random biopsy | Last biopsy |
| All biopsies | 1.12 (1.03-1.22) | 1.20 (1.13-1.27) | 1.29 (1.23-1.35) | 1.18 (1.15-1.21) | 1.20 (1.17-1.22) | 1.21 (1.18-1.23) | 1.18 (1.12-1.24) | 1.18 (1.12-1.24) | 1.21 (1.15-1.26) |
| No rejection | 1.18 (1.09-1.27) | 1.20 (1.12-1.28) | 1.23 (1.16-1.31) | 1.14 (1.10-1.18) | 1.14 (1.11-1.18) | 1.14 (1.11-1.18) | 1.14 (0.98-1.33) | 1.12 (0.95-1.32) | 1.16 (0.99-1.36) |
| All but No rejection | 1.13 (1.04-1.23) | 1.16 (1.1-1.24) | 1.24 (1.17-1.31) | 1.17 (1.14-1.20) | 1.16 (1.13-1.19) | 1.18 (1.15-1.21) | 1.14 (1.08-1.20) | 1.16 (1.10-1.22) | 1.19 (1.13-1.26) |
| AMR and/or TCMR | 1.11 (1.01-1.22) | 1.19 (1.11-1.27) | 1.20 (1.13-1.28) | 1.16 (1.12-1.20) | 1.16 (1.12-1.20) | 1.15 (1.12-1.19) | 1.08 (1.02-1.14) | 1.11 (1.04-1.17) | 1.12 (1.05-1.19) |
| TCMR | 1.10 (1.00-1.22) | 1.11 (1.02-1.21) | 1.16 (1.07-1.25) | 1.16 (1.11-1.21) | 1.14 (1.10-1.19) | 1.13 (1.09-1.18) | 1.06 (0.99-1.15) | 1.10 (1.02-1.18) | 1.11 (1.03-1.19) |
| (Borderline) TCMR | 1.14 (1.05-1.25) | 1.23 (1.15-1.33) | 1.23 (1.15-1.32) | 1.17 (1.13-1.21) | 1.16 (1.12-1.20) | 1.16 (1.13-1.20) | 1.12 (1.05-1.18) | 1.15 (1.08-1.21) | 1.16 (1.09-1.22) |
| PVAN | 1.25 (1.05-1.49) | 1.25 (1.05-1.50) | 1.24 (1.06-1.46) | 1.16 (1.02-1.31) | 1.17 (1.03-1.32) | 1.17 (1.03-1.33) | 0.86 (0.59-1.24) | 0.85 (0.59-1.23) | 0.85 (0.60-1.22) |
| AMR | 1.14 (1.00-1.29) | 1.14 (1.03-1.25) | 1.19 (1.10-1.29) | 1.17 (1.13-1.22) | 1.18 (1.13-1.22) | 1.17 (1.12-1.21) | 1.08 (1.00-1.16) | 1.08 (1.01-1.17) | 1.10 (1.02-1.18) |
| Probable AMR /MVI/AMR | 1.11 (1.01-1.22) | 1.20 (1.12-1.30) | 1.22 (1.14-1.30) | 1.16 (1.13-1.20) | 1.17 (1.13-1.21) | 1.17 (1.13-1.20) | 1.09 (1.03-1.16) | 1.10 (1.04-1.17) | 1.14 (1.07-1.21) |
| Mixed rejection | 1.19 (1.01-1.41) | 1.21 (0.99-1.47) | 1.26 (1.05-1.50) | 1.15 (1.08-1.23) | 1.16 (1.09-1.24) | 1.14 (1.08-1.22) | 1.07 (0.98-1.18) | 1.07 (0.97-1.17) | 1.07 (0.97-1.17) |

µ

**Table S11 Hazard ratios (with 95% confidence interval) of the activity and chronicity indices, adjusted for time-post-transplantation, in the subcategories of AMR.**

| | Activity index | | | | Chronicity index | | |
|---|---|---|---|---|---|---|---|
| | *First biopsy* | *Random biopsy* | *Last biopsy* | | *First biopsy* | *Random biopsy* | *Last biopsy* |
| **all AMR** | 1.11 (1.08-1.15) | 1.10 (1.07-1.14) | 1.11 (1.07-1.14) | **all AMR** | 1.14 (1.10-1.17) | 1.14 (1.10-1.17) | 1.16 (1.12-1.19) |
| **active AMR** | 1.12 (1.08-1.17) | 1.14 (1.10-1.19) | 1.13 (1.09-1.18) | **active AMR** | 1.09 (1.02-1.17) | 1.13 (1.06-1.20) | 1.12 (1.06-1.20) |
| **chronic active AMR** | 1.11 (1.06-1.17) | 1.11 (1.05-1.16) | 1.12 (1.07-1.18) | **chronic active AMR** | 1.16 (1.10-1.23) | 1.17 (1.10-1.24) | 1.17 (1.10-1.24) |
| **chronic AMR** | 1.14 (1.00-1.31) | 1.14 (0.99-1.31) | 1.13 (0.99-1.30) | **chronic AMR** | 1.15 (1.04-1.28) | 1.15 (1.03-1.28) | 1.15 (1.03-1.28) |

**TABLE S12 Missingness among the lesions scores per cohort as absolute numbers (and percentage).**

This table also includes potential candidate lesions for the development of the indices.

| Cohort | Derivation N= 6272 | European validation N=11043 | US validation N=2185 |
|---|---|---|---|
| **g** | 6 (0.1) | 27 (0.2) | 5 (0.2) |
| **ptc** | 250 (4.0) | 43 (0.4) | 10 (0.5) |
| **t** | 2 (0.0) | 20 (0.2) | 3 (0.1) |
| **i** | 2 (0.0) | 14 (0.1) | 3 (0.1) |
| **v** | 11 (0.2) | 452 (4.1) | 17 (0.8) |
| **C4d_PTC** | 55 (0.9) | 891 (8.1) | 12 (0.5) |
| **thrombi** | 8 (0.1) | 3891 (35.2) | 2185 (100.0) |
| **cg** | 10 (0.2) | 283 (2.6) | 13 (0.6) |
| **ci** | 6 (0.1) | 74 (0.7) | 7 (0.3) |
| **ct** | 4 (0.1) | 74 (0.7) | 7 (0.3) |
| **cv** | 10 (0.2) | 538 (4.9) | 28 (1.3) |
| **ah** | 6 (0.1) | 1635 (14.8) | 1951 (89.3) |
| **mm** | 6 (0.1) | 5493 (49.7) | 2185 (100.0) |
| **gs** | 92 (1.5) | 1613 (14.6) | 1950 (89.2) |

**Figure S1 Distribution of the activity index (panel A), chronicity index (panel B), AMR index (panel C) and TCMR index (panel D) per indication status in the derivation and validation cohort.** Biopsies with unknown indication/protocol status were excluded for this analysis. All comparisons protocol vs indication had with Student t-test p-values lesser to 0.001.

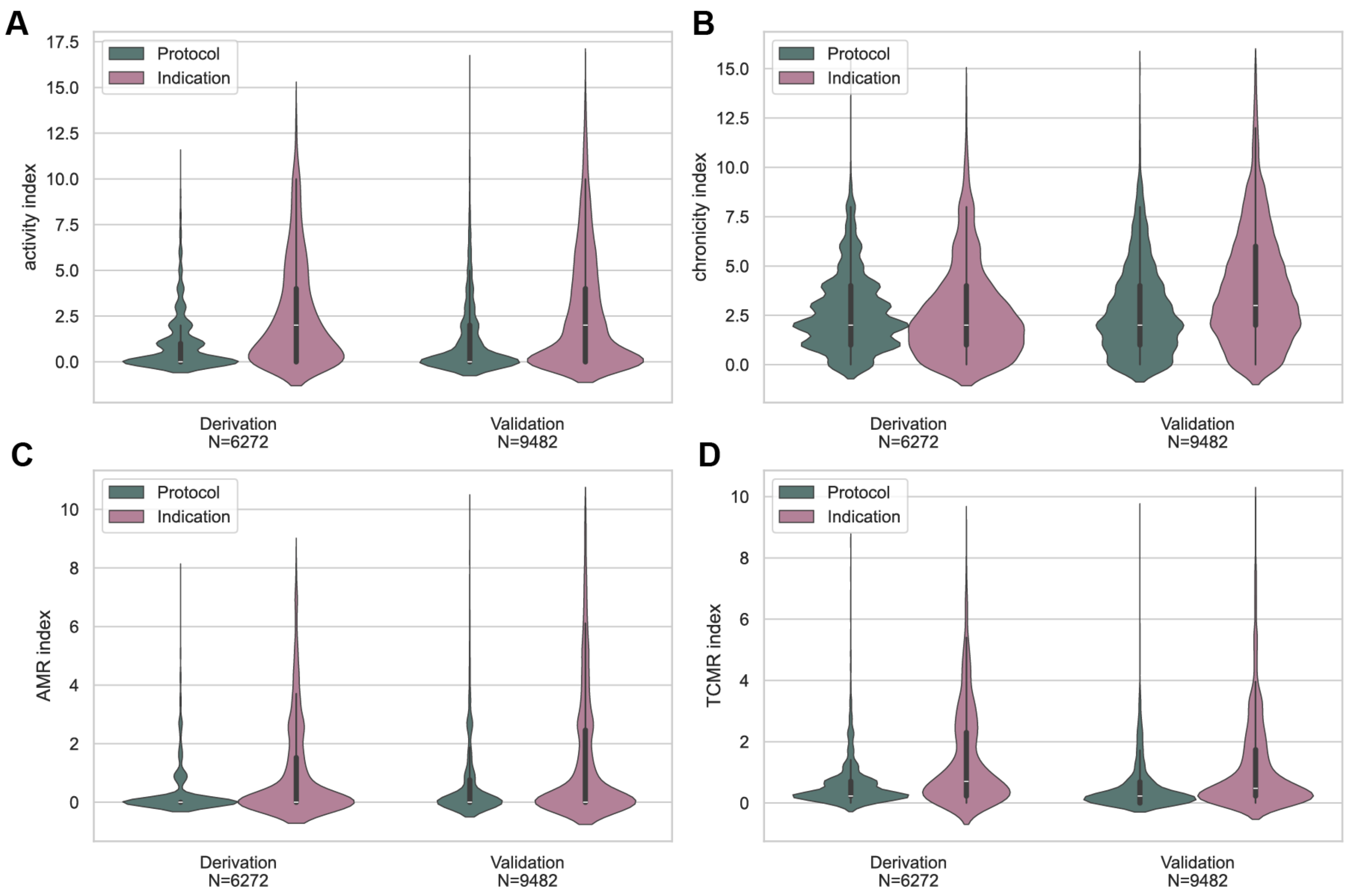

**Figure S2 Distribution of activity and chronicity indices in AMR and TCMR subcategories**

**A** Distribution of the activity and chronicity indices across the subcategories of AMR (N=1715, whole cohort). All Student t-test pairwise comparisons have a p-values< 0.001. However, the subcategories of AMR exhibit significant overlap in both indices. For instance, some chronic AMR biopsies have higher activity index than some active AMR.

**B** Distribution of the activity and chronicity indices across the subcategories of TCMR, on a subset of the derivation cohort for which critical Banff lesions (ti, i-IFTA) for the diagnostic of chronic active TCMR (caAMR) were available (N=1635). 42 Cases were caAMR (2.6%), out of which 10 were diagnosed with concomitant acute TCMR. All Student t-test pairwise comparison have a p-values< 0.001, except for the comparison of the chronicity index between chronic active TCMR vs acute and caTCMR: p-value= 0.74. Note the significant overlaps of indices between the subcategories.

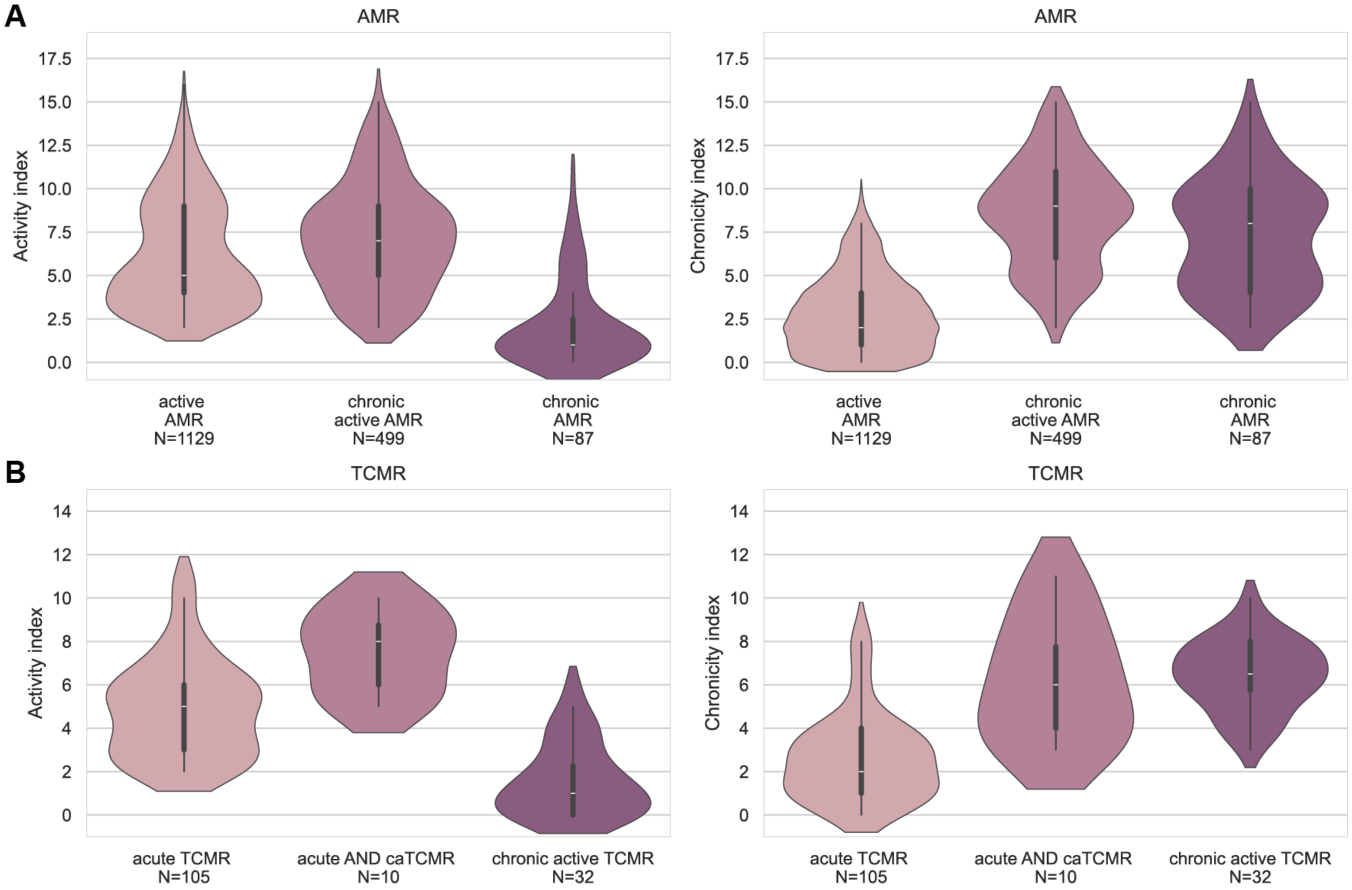

**Figure S3 Distribution of the four indices in the European validation cohort (N= 10339 biopsies)**

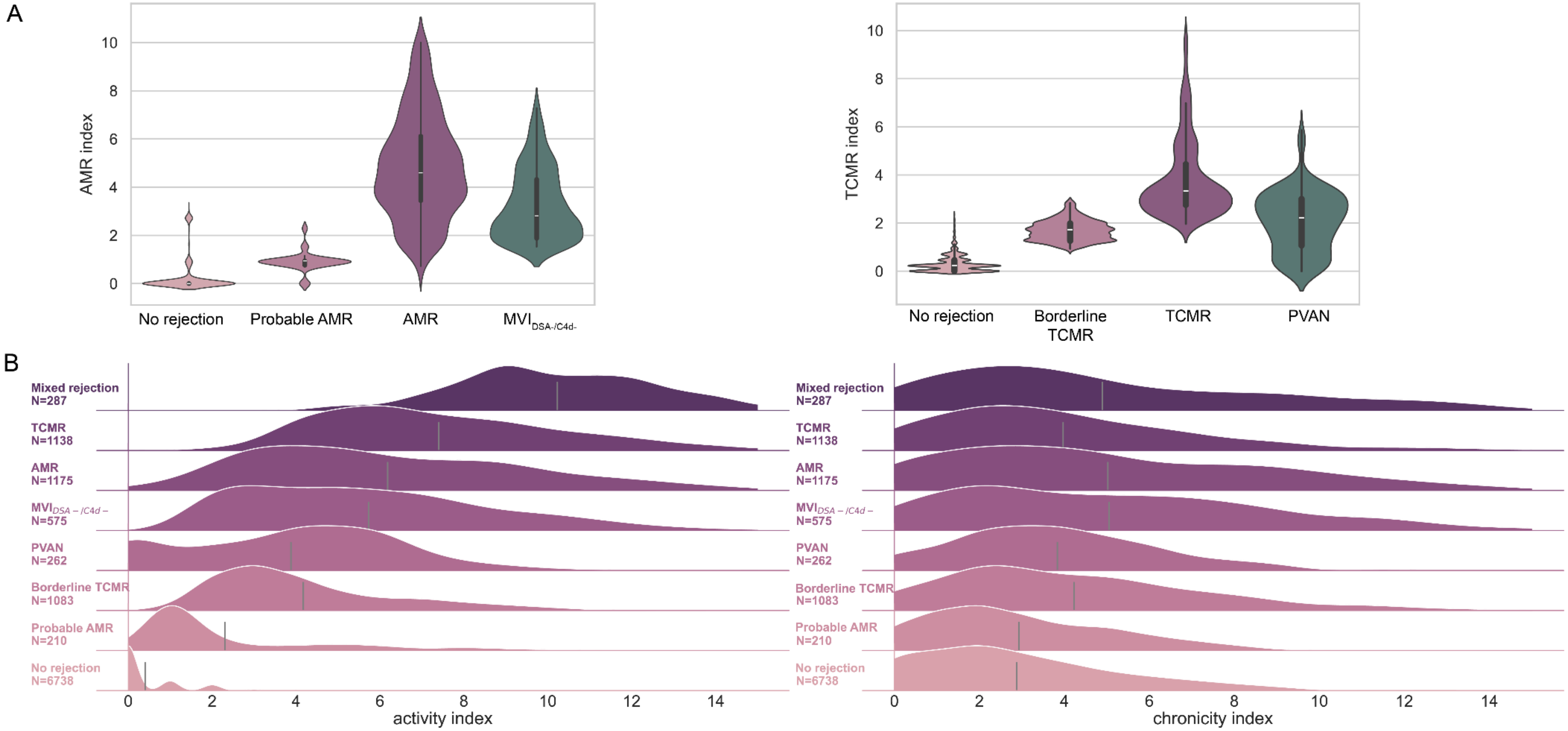

**Figure S4 Distribution of the four indices in the US validation cohort (N=2162 biopsies)**

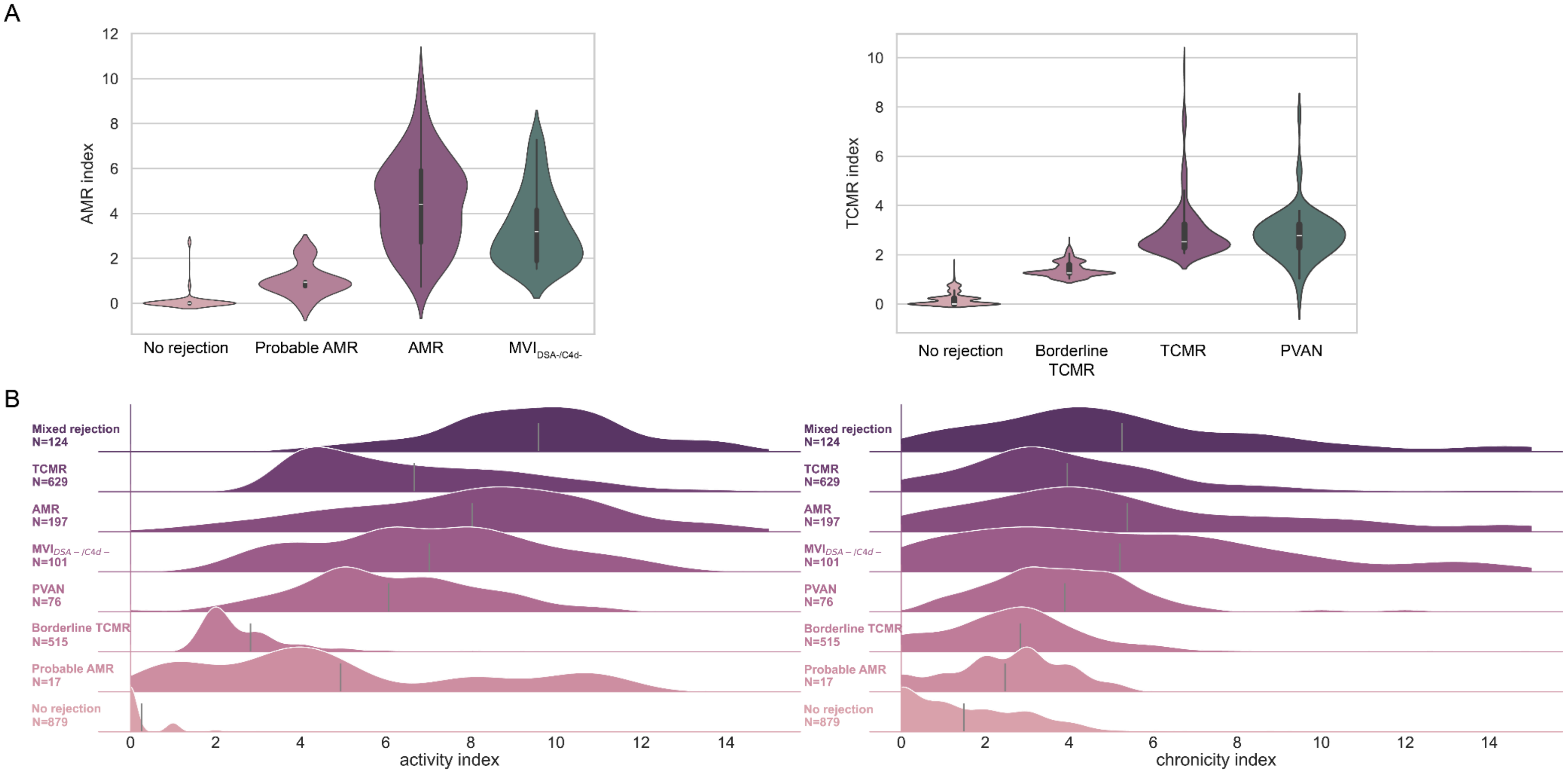

**Figure S5 Decision curve analyses for the prediction of AMR (panel A) and TCMR (panel B) per cohort.** The relevant indices were turned into probabilities based on logistic regression models. Their predictive performance was compared to models 2 (without HLA-DSA and PRA) from Sikosana et al.

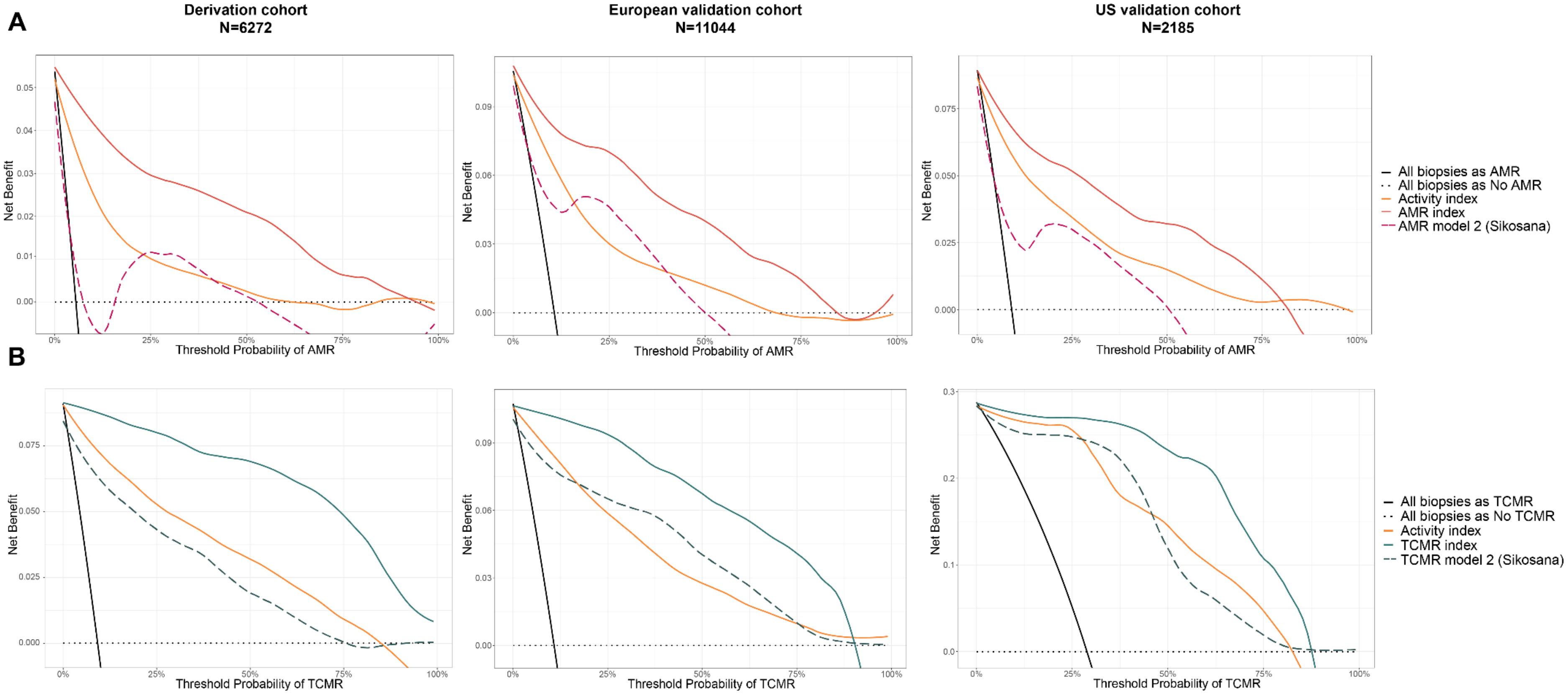

**Figure S6 Kaplan-Meier curves of the AMR-related (Panel A) and TCMR-related (Panel C) Banff diagnostic categories and stratification of the AMR (panel B) and TCMR (panel D) indices in three strata based on arbitrary cut-offs (all cohorts, N=19500).** *No AMR* refers to all biopsies that are not AMR, Probable AMR or $MVI_{DSA-/C4d-}$. Similarly, *No TCMR* refers to all biopsies that are not TCMR or Borderline TCMR. Due to the negative exponential aspect of the index distributions, the discretization was based on the index value at the $75^{th}$ and $95^{th}$ percentiles, corresponding to the following pairs of thresholds: 0.9 and 4.6 and; 1.3 and 3.3, and for the ARM index and TCMR index, respectively. The Kaplan-Meier curves use the last biopsy per individual. P-values refer to log-rank tests.

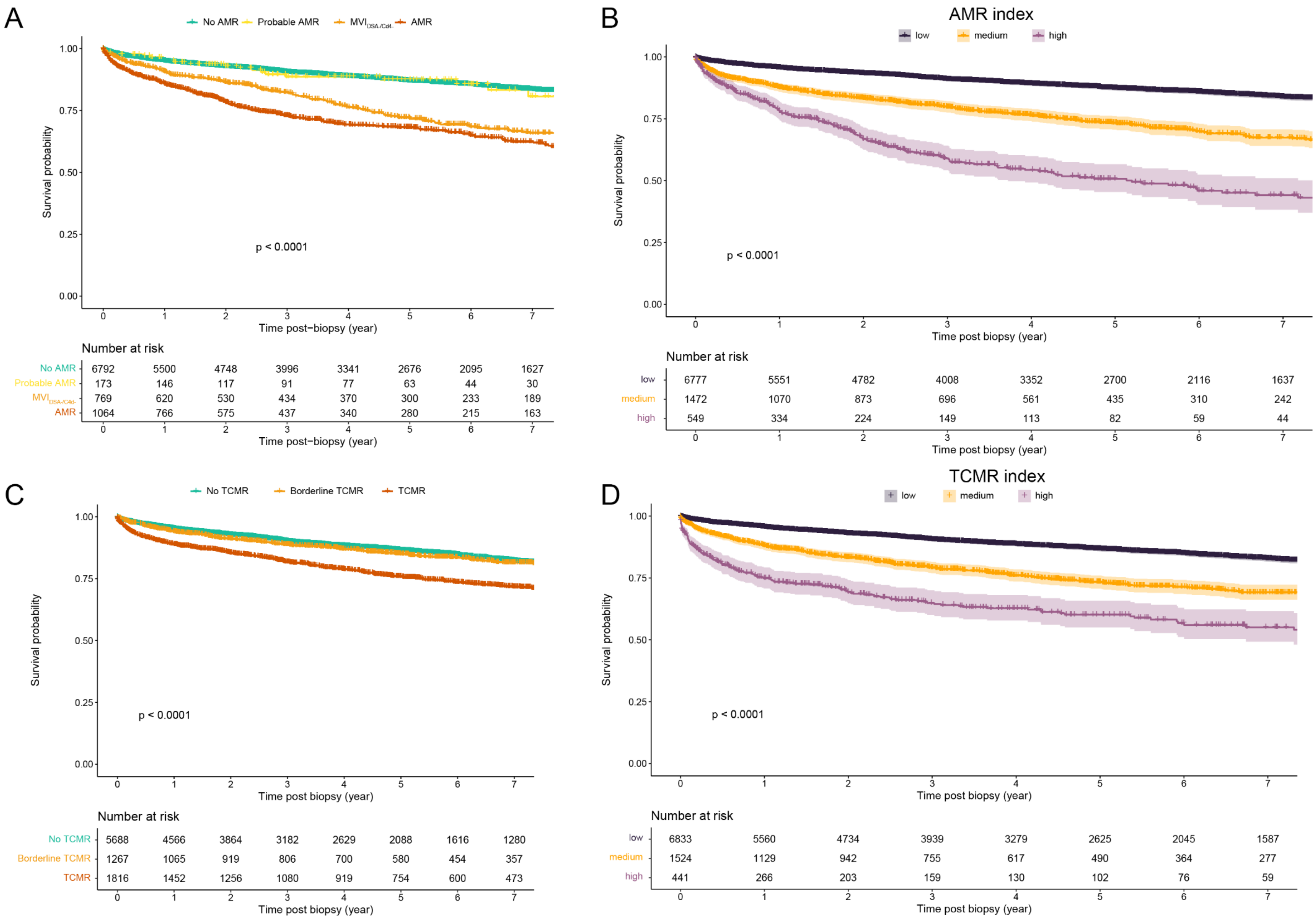
A
No AMR
Probable AMR
$MVI_{DSA-/C4d-}$
AMR
Survival probability
1.00
0.75
0.50
0.25
0.00
p < 0.0001
Time post−biopsy (year)
0 1 2 3 4 5 6 7
Number at risk
No AMR 6792 5500 4748 3996 3341 2676 2095 1627
Probable AMR 173 146 117 91 77 63 44 30
$MVI_{DSA-/C4d-}$ 769 620 530 434 370 300 233 189
AMR 1064 766 575 437 340 280 215 163
B
AMR index
low
medium
high
Survival probability
1.00
0.75
0.50
0.25
0.00
p < 0.0001
Time post biopsy (year)
0 1 2 3 4 5 6 7
Number at risk
low 6777 5551 4782 4008 3352 2700 2116 1637
medium 1472 1070 873 696 561 435 310 242
high 549 334 224 149 113 82 59 44
C
No TCMR
Borderline TCMR
TCMR
Survival probability
1.00
0.75
0.50
0.25
0.00
p < 0.0001
Time post biopsy (year)
0 1 2 3 4 5 6 7
Number at risk
No TCMR 5688 4566 3864 3182 2629 2088 1616 1280
Borderline TCMR 1267 1065 919 806 700 580 454 357
TCMR 1816 1452 1256 1080 919 754 600 473
D
TCMR index
low
medium
high
Survival probability
1.00
0.75
0.50
0.25
0.00
p < 0.0001
Time post biopsy (year)
0 1 2 3 4 5 6 7
Number at risk
low 6833 5560 4734 3939 3279 2625 2045 1587
medium 1524 1129 942 755 617 490 364 277
high 441 266 203 159 130 102 76 59

**Figure S7 Stratification of the AMR (panel A) and $MVI_{DSA-/C4d-}$ (panel C) cases based on the AMR index and TCMR (panel B) and Borderline TCMR (panel D) cases based on TCMR index with arbitrary cut-offs (all cohorts, N=19500) and corresponding survival trajectory (Kaplan-Meier curves).** The two arbitrary cut-off values were selected to divide the index (min-max) range of each Banff category into 3 levels of same interval: Moderate (strictly below 1st cut-off), High (higher or equal to 1st cut-off, strictly lower than 2nd cut off) and Severe (strictly higher than 2nd cut-off). The corresponding pair of cut-offs are as follows: AMR: 3.8, 6.9; $MVI_{DSA-/C4d-}$: 3.4, 5.4; TCMR: 4.6, 7.2; Borderline TCMR: 1.6, 2.2. The Kaplan-Meier curves use the last biopsy per individual in case of repeated biopsies. P-values refer to log-rank tests.

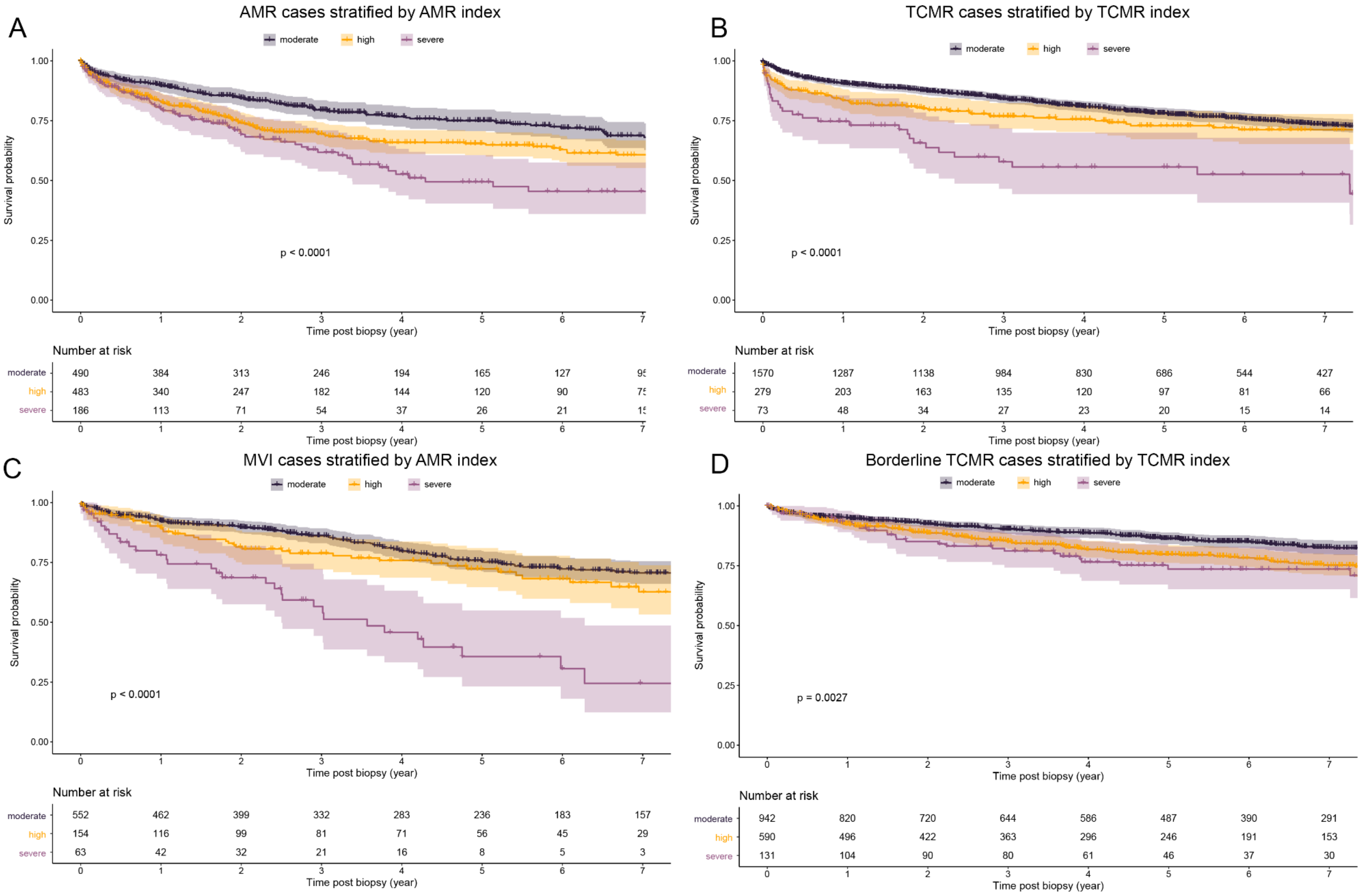

**Figure S8 Association with graft failure for both versions of the activity index, using either the lesion i or ti.**

Based on a subset of the derivation cohort with both available lesions (N=3724). Replacing i with ti in the activity index does not modify the association of the index with graft failure in the different diagnostic categories.

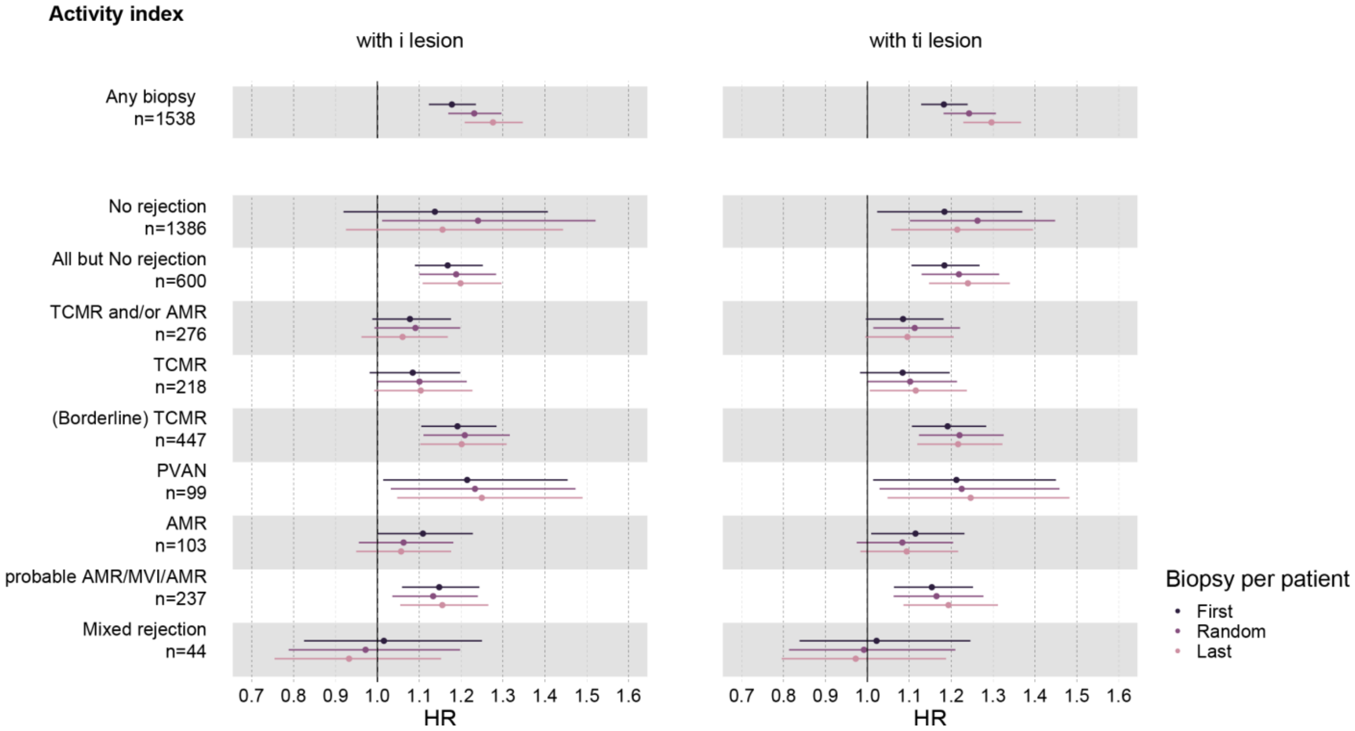

**Figure S9 Linear association of activity and chronicity indices with risk of graft failure.** Both activity and chronicity indices linearly associate with graft outcome (p-value for non-linear effect with restricted cubic splines with 3 knots: 0.86 and 0.83, respectively). Models based on a random biopsy per patient, all cohorts (N=19,500)

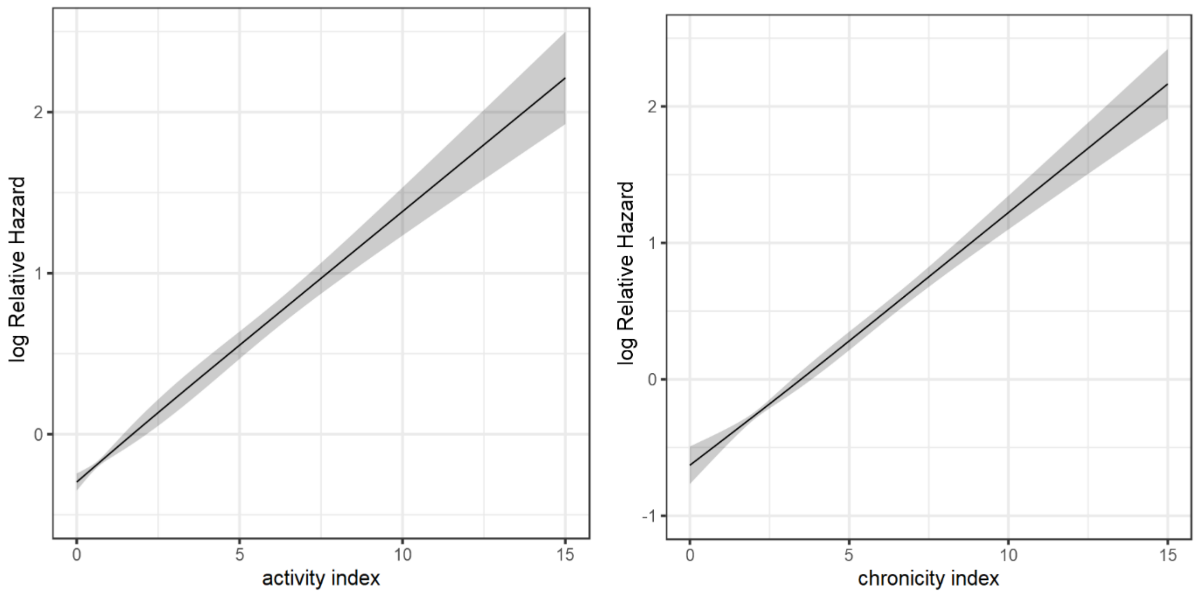

**Figure S10 Sensitivity analyses: hazard ratios (with 95% CI) of the activity and chronicity indices with graft failure per biopsy indication status (indication vs protocol), adjusted for time-post-transplantation.**

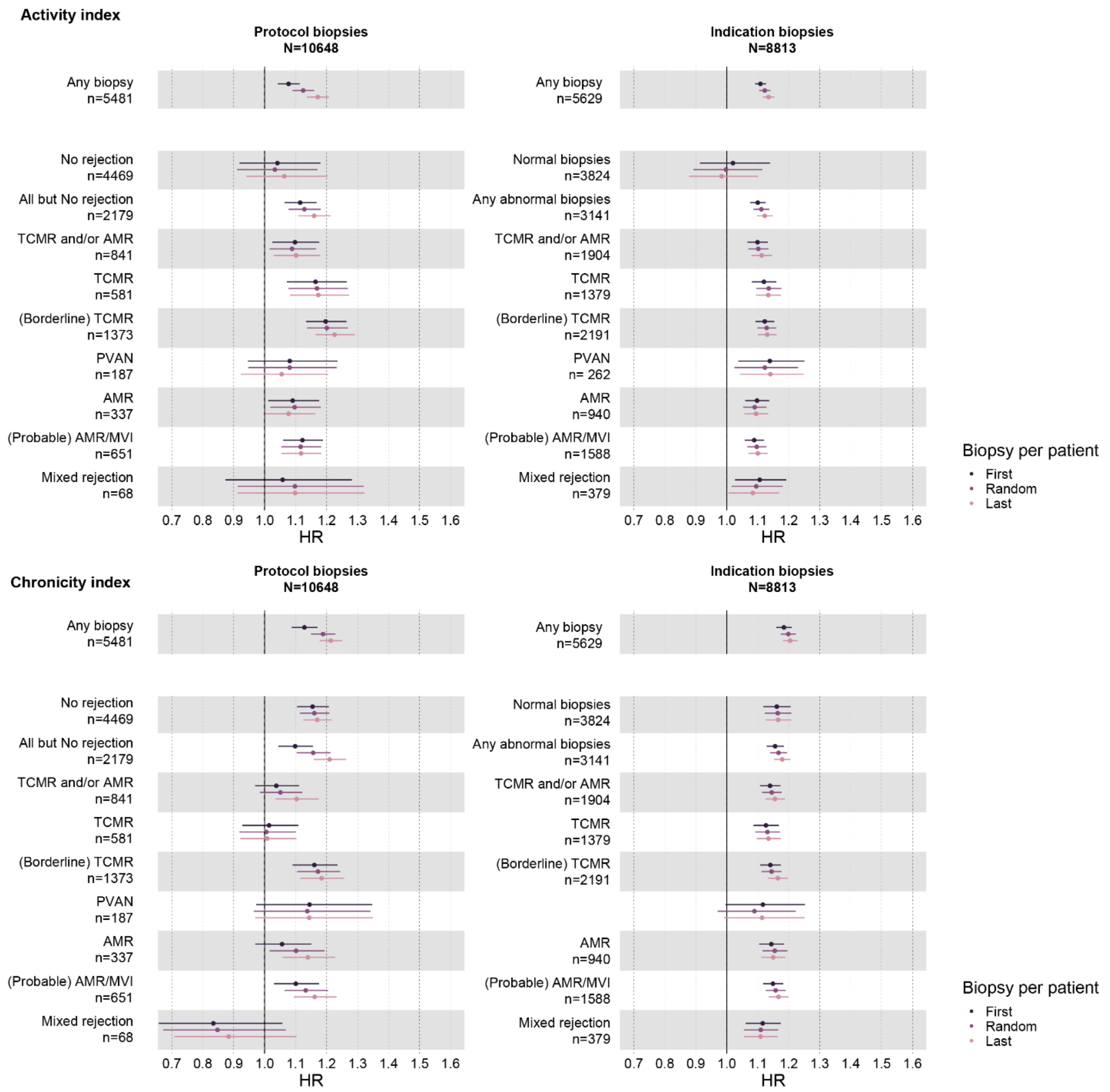

**Figure S11 Hazard ratios (with 95% confidence interval) of the activity and chronicity indices, adjusted for time-post-transplantation, in the subcategories of AMR.**

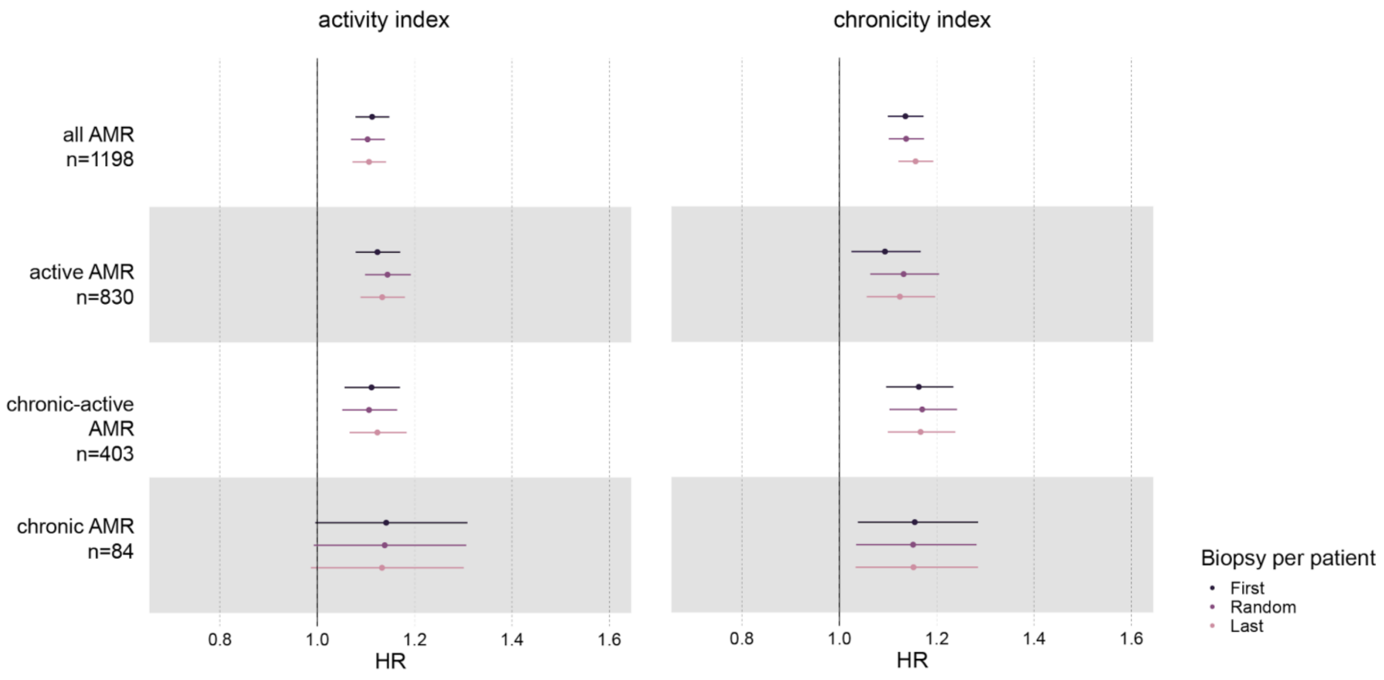
